# Histologically validated diffusion MRI signatures of neuroinflammation and neurodegeneration in Alzheimer disease

**DOI:** 10.64898/2026.06.11.26354743

**Authors:** Qing Wang, Meng Jiang, Aime Luna, Xuan Niu, Yuan Nan, Qiuchang Sun, Richard J. Perrin, Erin Franklin, Carlos Cruchaga, Shaney Flores, Tammie L.S. Benzinger, Yong Wang

**Author notes:** (Send correspondence to: Qing Wang, Ph.D. Mallinckrodt Institute of Radiology,Washington University in St. Louis, School of Medicine 4525 Scott Ave., Campus Box 8225, St. Louis, MO 63110.

## Abstract

Noninvasive neuroinflammation measurement remains a major barrier for Alzheimer disease (AD) therapeutics. We present generalized diffusion basis spectrum imaging (g-DBSI), a diffusion MRI framework that decomposes the tissue signal into biologically interpretable microstructural compartments. In postmortem Knight ADRC brains, g-DBSI-derived restricted isotropic fraction (RIF) and restricted anisotropic fraction (RAF) mapped cellularity and neurofilament density, while their ratio (RIF/RAF) tracked inflammatory cell density and peri-plaque amyloid-beta with higher specificity and regional consistency than RIF alone. In 112 living Knight ADRC participants stratified by PET amyloid, g-DBSI metrics showed amyloid-dependent trajectories: in low-amyloid individuals, RIF and RAF rose together with amyloid, consistent with early neuropil expansion and glial elaboration, whereas in high-amyloid individuals, RIF/RAF increased, and RAF declined, indicating established neuroinflammatory remodeling and neurofilament loss. CSF proteomics linked RIF/RAF to glia-enriched immune and vascular pathways, supporting g-DBSI as a clinically compatible MRI biomarker of neuroinflammation and neurodegeneration in AD.

**Teaser:** g-DBSI provides noninvasive MRI biomarkers of neuroinflammation and neurodegeneration in AD, validated by histopathology and CSF proteomics.

## Introduction

Neuroinflammation is a fundamental driver and modifier of neurodegenerative disease biology and therapeutic response. Across Alzheimer disease (AD) and related disorders, microglia and astrocytes shape synaptic integrity (*1*), axonal health (*2*), edema (*3*), and amyloid plaque dynamics (*4*), influencing both disease progression and treatment-related side-effects such as amyloid-related imaging abnormalities (ARIA) (*5, 6*). Yet quantifying glial activation, edema, and axonal injury with specificity and regional detail remains challenging in routine practice and clinical trials. Established diffusion tensor imaging (DTI) metrics (e.g., fractional anisotropy [FA], mean diffusivity [MD]) are sensitive but nonspecific, as they are confounded by edema, cellular proliferation, and axonal changes. Molecular positron emission tomography (PET) offers cell-type- or pathway-specific imaging but is constrained by tracer availability, off-target binding, limited spatial resolution, radiation exposure, and cost, which limit its use for longitudinal monitoring (*7–10*).

Advanced diffusion MRI methods offer distinct and complementary perspectives on tissue microstructure, each emphasizing specific features of neuroinflammation through its underlying modeling framework. Free-water imaging (FWI) (*11*) separates an extracellular isotropic component associated with increased free-water content, thereby providing sensitivity to edema-related inflammatory changes. Neurite Orientation Dispersion and Density Imaging (NODDI) (*12*) characterizes neurite density and orientation dispersion using a three-compartment model that estimates the intracellular volume fraction (ICVF), the orientation dispersion index (ODI), and the isotropic free-water fraction (ISOVF). Within this framework, increases in the ISOVF can reflect elevated free-water content, such as vasogenic or inflammatory edema, providing a complementary perspective on tissue water and microstructural changes that may accompany neuroinflammatory processes, even though NODDI does not specifically differentiate edema related to reactive glia from other sources of extracellular free water. Soma and neurite density imaging (SANDI) model (*13*) separates water diffusion in cell bodies (somas) from that in neurites, providing *in vivo* estimates of soma and neurite density and size across brain tissue. In the context of neuroinflammation, SANDI can be used to infer microglial and astrocytic activation and associated neurite pathology by detecting increased soma density and altered neurite density/dispersion, requiring high b-values and strong gradients. Diffusion kurtosis imaging (DKI) (*14*) captures deviations from Gaussian diffusion, reflecting microstructural complexity through higher-order signal behavior. More detailed biophysical models (e.g., CHARMED/AxCaliber (*15*), spherical mean technique (*16*), VERDICT/RSI (*17*), Mean apparent propagator MRI (*18*)) can improve sensitivity to restriction or orientation dispersion under specific geometric assumptions.

In parallel, diffusion basis spectrum imaging (DBSI) is an advanced diffusion MRI framework that decomposes the diffusion signal into multiple anisotropic and isotropic components, enabling in vivo estimation of axonal injury, demyelination, and, in particular, inflammation (e.g., cellular infiltration and vasogenic edema) (*19, 20*). Generalized-DBSI (g-DBSI), developed in this study, extends this model from its original white matter (WM) focus to the cortex to better capture the more isotropic, microstructurally heterogeneous cortical microenvironment while preserving sensitivity to inflammatory processes. g-DBSI decomposes the diffusion signal into a continuous spectrum of anisotropic (fiber) and isotropic (cellular/edematous) components and estimates biologically interpretable fractions: restricted isotropic fraction (RIF), hindered isotropic fraction (HIF), restricted anisotropic fraction (RAF), hindered anisotropic fraction (HAF), and perfusion fraction (PF). A RIF/RAF ratio may serve as a potential neuroinflammation marker by emphasizing glial-associated restricted diffusion while discounting free-water effects. This spectrum-based modeling is inherently tolerant of crossing fibers and mixed tissues, yielding quantitative maps that are comparable across regions, individuals, and time points.

Despite the promising ability of the g-DBSI framework to disentangle coexisting inflammatory and neurodegenerative processes, two hurdles have limited broad clinical translation: (i) definitive validation against gold-standard histopathology across diverse human brain regions, and (ii) molecular anchoring of g-DBSI metrics to circulating biomarkers that report on glial states. In this study, to address these gaps, we combined *ex situ* multi-b-value diffusion MRI of postmortem human brain tissue blocks with quantitative histology (hematoxylin and eosin histochemistry [H&E] for cellularity; 2F11 for neurofilaments; GFAP for astrocytes, IBA-1 for microglia; and 10D5 for amyloid-beta peptide deposits). After precise registration between MRI and histology, we evaluated whether g-DBSI-derived metrics correlate with histologically quantified cellularity, neurofilaments, and inflammatory cell markers in the hippocampus, striatum, and multiple cortical regions. In parallel, we tested whether the whole-cortex g-DBSI inflammation index is associated with inflammatory signatures in cerebrospinal fluid (CSF) proteomics. This integrated imaging-histology-proteomics framework establishes biological specificity for g-DBSI metrics.

## Results

### g-DBSI-derived restricted isotropic fraction tracks histological cellularity

Forty-two formalin-fixed postmortem tissue blocks, representing seven brain regions vulnerable to AD pathology, were obtained from six Charles F. and Joanne Knight Alzheimer’s Disease Research Center (Knight ADRC) participants. Participant demographics are summarized in **Table S1**. The seven brain regions include: (i) MFG: middle frontal gyrus, (ii) ACG: anterior cingulate gyrus, (iii) Striatum: striatum with nucleus accumbens and olfactory cortex, (iv) HPC: hippocampus with parahippocampal, fusiform, and inferior temporal gyri, (v) PCG/PCu: posterior cingulate gyrus and precuneus, (vi) PL: parietal lobe (angular gyrus), and (vii) OL: occipital lobe (calcarine sulcus and parastriate cortex) (**Fig. S1**). All specimens underwent *ex situ* g-DBSI scans, H&E staining, and immunohistochemistry (IHC) staining with primary antibodies of 2F11, GFAP, IBA-1, and 10DF to enable quantification of the total cellularity, neurofilaments, astrocytes, microglia, and amyloid deposition, respectively (**Fig. 1**). For each block, g-DBSI metric maps and quantitative histology-derived density maps were generated and aligned into a common voxel grid for region-wise spatial correlation analyses. Three blocks were excluded from this study due to broken histology slides and poor registration. Representative postmortem tissue blocks, H&E sections, H&E-based cell density maps, and corresponding g-DBSI RIF maps are shown in **Fig. 2A to 2B**. Within each sampled region, RIF was positively associated with H&E-derived cell density (**Fig. 2Bd**; all *p* < 0.001), and these relationships remained significant after adjustment for postmortem interval (PMI: time between death and brain tissue fixation). The mean correlation coefficient across all sampled regions was 0.71 (SD = 0.11, n =39).

**Figure 1:**
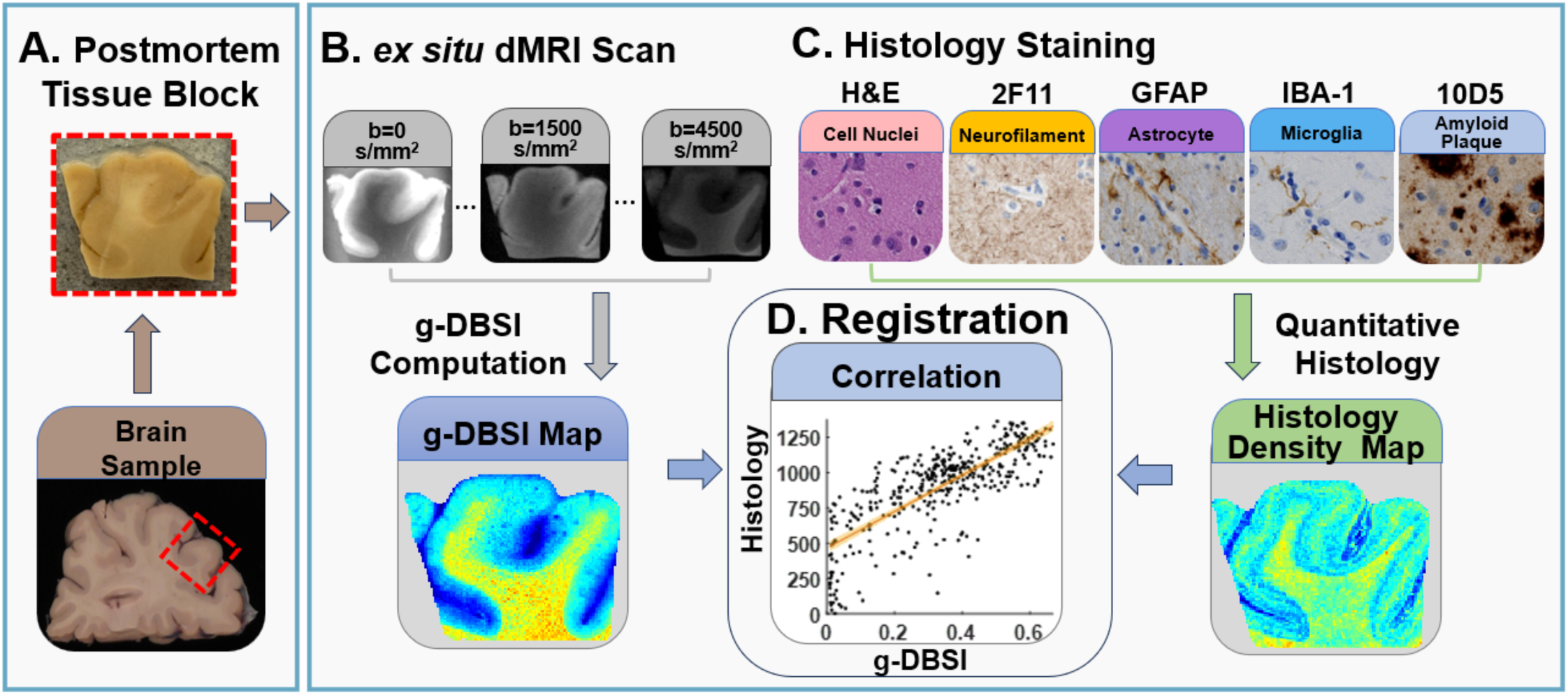
Workflow for *ex situ* validation of g-DBSI microstructural metrics using quantitative histology. (**A**) Postmortem human brain samples were obtained, and a cortical or subcortical tissue block was dissected from each region of interest (red dashed box) for paired imaging and histological analysis. (**B**) Each tissue block underwent high-resolution *ex situ* diffusion MRI acquired across multiple b-values (b = 0, 1500, and 4500 s/mm²), from which generalized diffusion basis spectrum imaging (g-DBSI) was computed to generate quantitative microstructural parameter maps. (**C**) Adjacent sections from the same tissue block were processed for histology and immunohistochemistry (IHC), including H&E (cell nuclei/cellularity), 2F11 (neurofilament/axonal integrity), GFAP (astrocytes), IBA-1 (microglia), and 10D5 (amyloid plaques). Each stain was converted to a quantitative histology density map using automated image analysis. (**D**) g-DBSI parameter maps were spatially co-registered to the corresponding histology density maps, and region-wise spatial correlation analyses were performed to quantitatively assess the correspondence between g-DBSI-derived microstructural metrics and histopathological markers.

**Figure 2:**
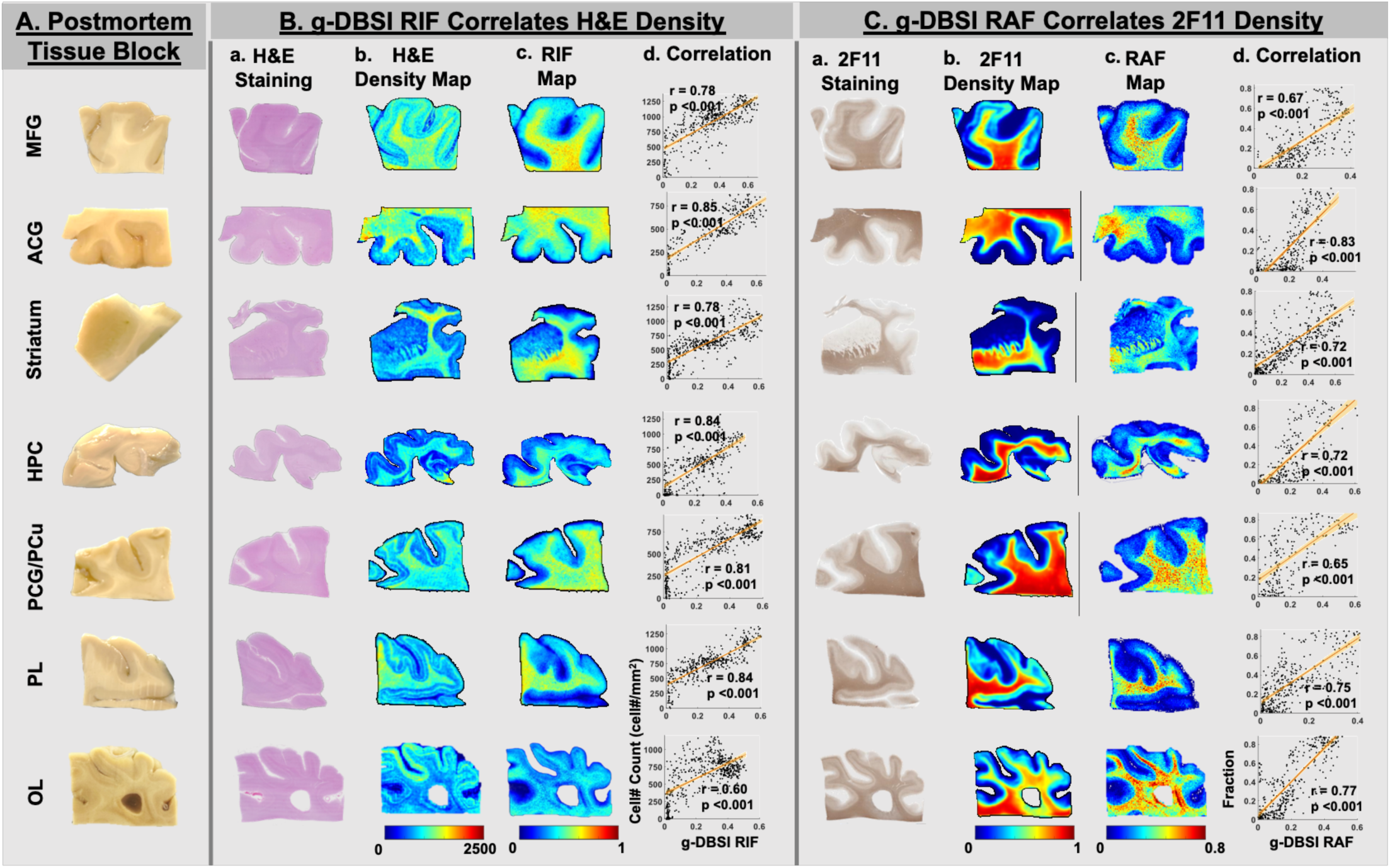
g-DBSI-derived RIF and RAF correlate with quantitative cellularity and neurofilament histopathology density in postmortem human brain tissues. (**A**) Representative postmortem tissue blocks were obtained from multiple cortical and subcortical regions, including the middle frontal gyrus (MFG), parietal lobe (PL), hippocampus (HPC), striatum, anterior cingulate gyrus (ACG), posterior cingulate gyrus/precuneus (PCG/PCu), and occipital lobe (OL). (**B**) Histological validation of g-DBSI-derived restricted isotropic fraction (RIF) using H&E staining. For each region, H&E-stained sections (Ba) were converted to quantitative cell density maps (Bb) and co-registered to corresponding g-DBSI RIF maps (Bc). Spatial correlation analyses demonstrated significant positive correlations between g-DBSI RIF and H&E-derived cell density across all sampled regions (Bd; all *p* < 0.001). (**C**) Histological validation of g-DBSI-derived restricted anisotropic fraction (RAF) using neurofilament immunohistochemistry (2F11). For each region, 2F11-stained sections (Ca) were converted to quantitative neurofilament density maps (Cb) and co-registered to corresponding g-DBSI RAF maps (Cc). Spatial correlation analyses demonstrated significant positive correlations between g-DBSI RAF and 2F11-derived neurofilament density across all sampled regions (Cd; all *p* < 0.001). Color bars indicate relative cell density or neurofilament density (B-C, left panels) or g-DBSI fraction values (B-C, right panels).

### g-DBSI-derived restricted anisotropic fraction tracks neurofilament density

Across all brain regions, areas with elevated g-DBSI RAF on MRI colocalized with dense neurofilament immunoreactivity on 2F11 staining (**Fig. 2Ca-c**). RAF showed significant positive correlations with 2F11-derived neurofilament density in every specimen (**Fig. 2Cd**), capturing tract-like architecture in WM and anisotropic neurofilament-rich structures in gray matter (GM), while maintaining contrast in regions with mixed tissue composition. In GM, 2F11 labeling reflects neurofilament within neuronal cell bodies and dendritic processes, whereas in white matter it primarily indexes the density and integrity of myelinated axons; g-DBSI RAF was significantly associated with 2F11 across cortical and subcortical brain tissue blocks, and these associations persisted after accounting for potential confounds such as PMI. The mean correlation coefficient across all sampled regions was 0.59 (SD = 0.18, n =39).

### g-DBSI RIF/RAF correlates with the neuroinflammatory cell density

A map of the RIF/RAF ratio was generated for each postmortem brain tissue block. This ratio synthesizes two complementary measures into a single normalized index that quantifies the balance between inflammatory cellularity and organized neural tissue integrity, capturing the pathological signature of neuroinflammation: infiltration and proliferation of reactive astrocytes and microglia, and with concurrent loss or replacement of functional neural structures, which is a process that occurs across both gray and white matter compartments throughout the brain (*21*).

To investigate the relationship between the g-DBSI RIF/RAF ratio and neuroinflammatory cell density in AD, g-DBSI RIF/RAF maps were generated for each specimen and spatially registered to summed GFAP (astrocytes) and IBA-1 (microglia) histology density maps (**Fig. 3A-C**). WM and GM regions were segmented based on the dMRI B0 maps. Visually, RIF/RAF “hotspots” overlapped inflammatory cell-rich areas on IHC, and statistically, significant positive associations between RIF/RAF and histology-derived neuroinflammatory cell density were observed in all sampled regions in both WM and GM (all *p* < 0.001; **Fig. 3D**). These findings indicate that g-DBSI RIF/RAF ratio captures structurally relevant features of astrocytic and microglial activation.

**Figure 3.**
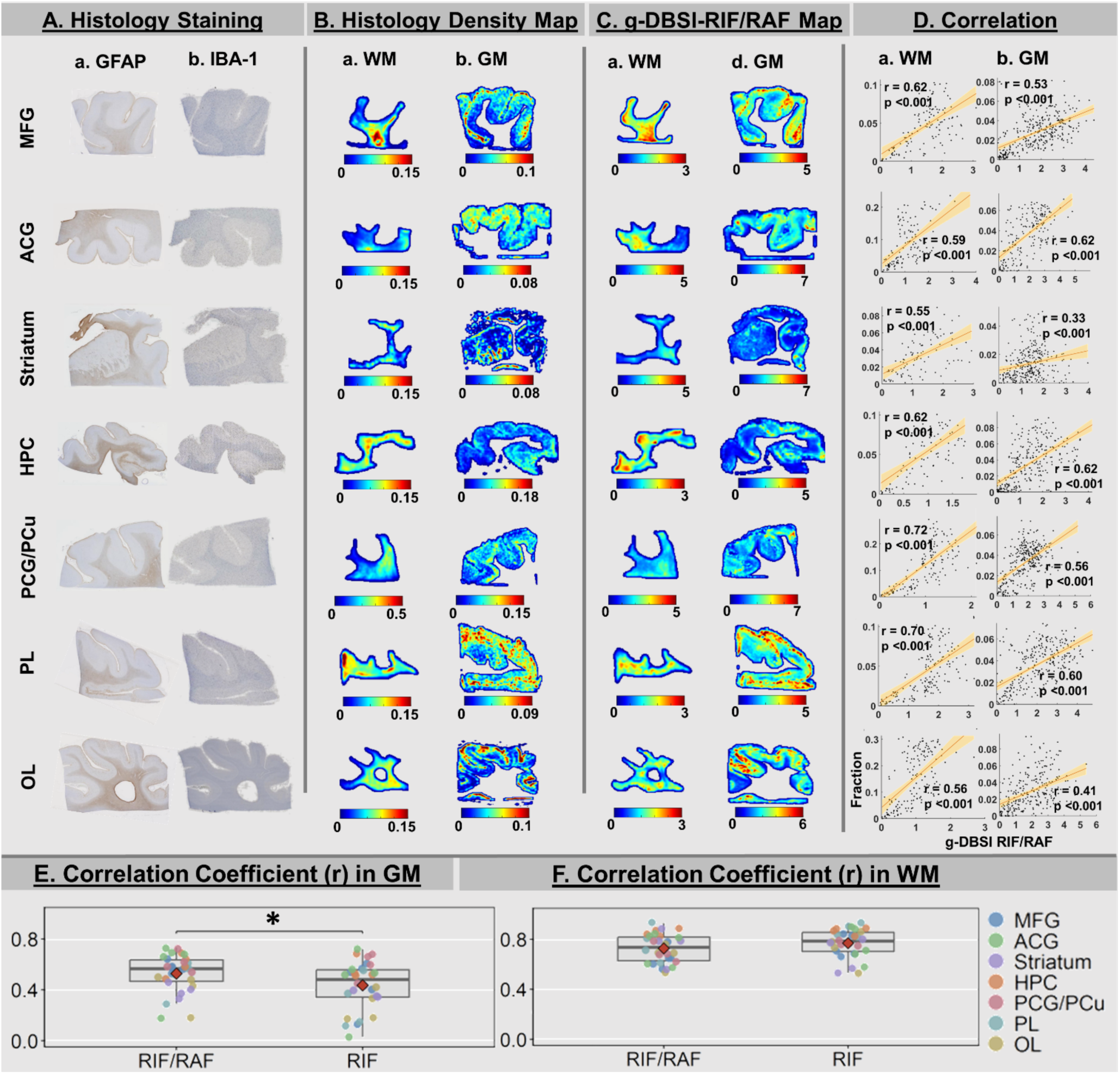
g-DBSI-derived RIF/RAF correlates with quantitative astrocytes and microglial histopathology density across brain regions in postmortem brain tissue blocks. (**A**) Representative GFAP (astrocytes) and IBA-1 (microglia) IHC staining slides from postmortem brain tissue blocks across seven brain regions: middle frontal gyrus (MFG), parietal lobe (PL), hippocampus (HPC), striatum, anterior cingulate gyrus (ACG), posterior cingulate/precuneus (PCG/PCu), and occipital lobe (OL). (**B**) GFAP/IBA-1 staining images were converted to quantitative histology density maps and segmented into white matter (WM) and gray matter (GM). (**C**) Corresponding g-DBSI-derived RIF/RAF maps were generated from the *ex situ* diffusion MRI data and segmented into WM and GM based on the b=0 image. (**D**) Spatial correlation analysis between g-DBSI RIF/RAF and combined GFAP/IBA-1 histology density demonstrated significant positive correlations in both WM and GM across all sampled regions (all *p* < 0.001). Color bars indicate relative histology density or g-DBSI RIF/RAF values. (**E–F**) Box plots summarizing region-wise Pearson correlation coefficients (r) between combined GFAP/IBA-1 histology density and RIF/RAF or RIF alone in GM and WM. Each colored dot represents one brain region. Boxes indicate the interquartile range (IQR); horizontal line, median; filled red diamond, mean; whiskers, 1.5×IQR; open circles, outliers by the 1.5×IQR rule. In GM, RIF/RAF showed significantly higher correlation with IBA1+GFAP density than RIF (\**p* = 0.013), whereas no significant difference was observed in WM (p = 0.153).

To further evaluate the specificity of the RAF normalization, correlations between g-DBSI RIF alone and inflammatory cell density maps were additionally computed in both WM and GM (**Fig. S2**). In representative cases, RIF exhibited weak spatial correlations with inflammatory cell density across several cortical and subcortical regions, including MFG, PL, and OL, suggesting that RIF alone is insufficient to capture local neuroinflammation without normalization to the anisotropic fiber signal. Consistent with this, the mean correlation coefficient for RIF/RAF was significantly higher than that for RIF in GM (**Fig. 3E**), whereas no significant difference between the two metrics was observed in WM (**Fig. 3F**). Moreover, the variance of the correlation coefficient was substantially lower for RIF/RAF than for RIF (0.11 vs. 0.19), demonstrating that RIF/RAF provides greater regional consistency than RIF alone in reflecting neuroinflammatory cell density across brain regions.

### *Ex situ* g-DBSI RIF/RAF spatially correlates with amyloid-beta deposition

To evaluate the spatial relationship between inflammation quantified by g-DBSI-derived RIF/RAF and amyloid-beta pathology stained by the 10D5, we compared g-DBSI RIF/RAF maps with 10D5 IHC density maps across seven postmortem brain regions (MFG, ACG, Striatum, HPC, PCG/PCu, PL, and OL) in five participants. The participant who has the Thal Stage of 1 was excluded in this analysis due to its low amyloid binding in the postmortem brain tissue blocks. Four representative regions (MFG, ACG, HPC, and PL) from one participant are shown in **Figure 4**. In each region, 10D5 staining intensity (**Fig. 4Ab-Db**) was calculated by the stained area divided by the voxel area. The corresponding g-DBSI RIF/RAF signal (**Fig. 4Ac-Dc**) exhibited visually concordant spatial distributions, with elevated signals co-localizing in cortical regions known to accumulate amyloid plaques. Quantitative region-wise correlation analyses confirmed significant positive associations between RIF/RAF and 10D5 density across all regions (**Fig. 4Ad-Dd**; all *p* < 0.001). The mean correlation coefficient across all postmortem brain tissues included in this analysis was 0.59 (SD = 0.12, n=32).

**Figure 4:**
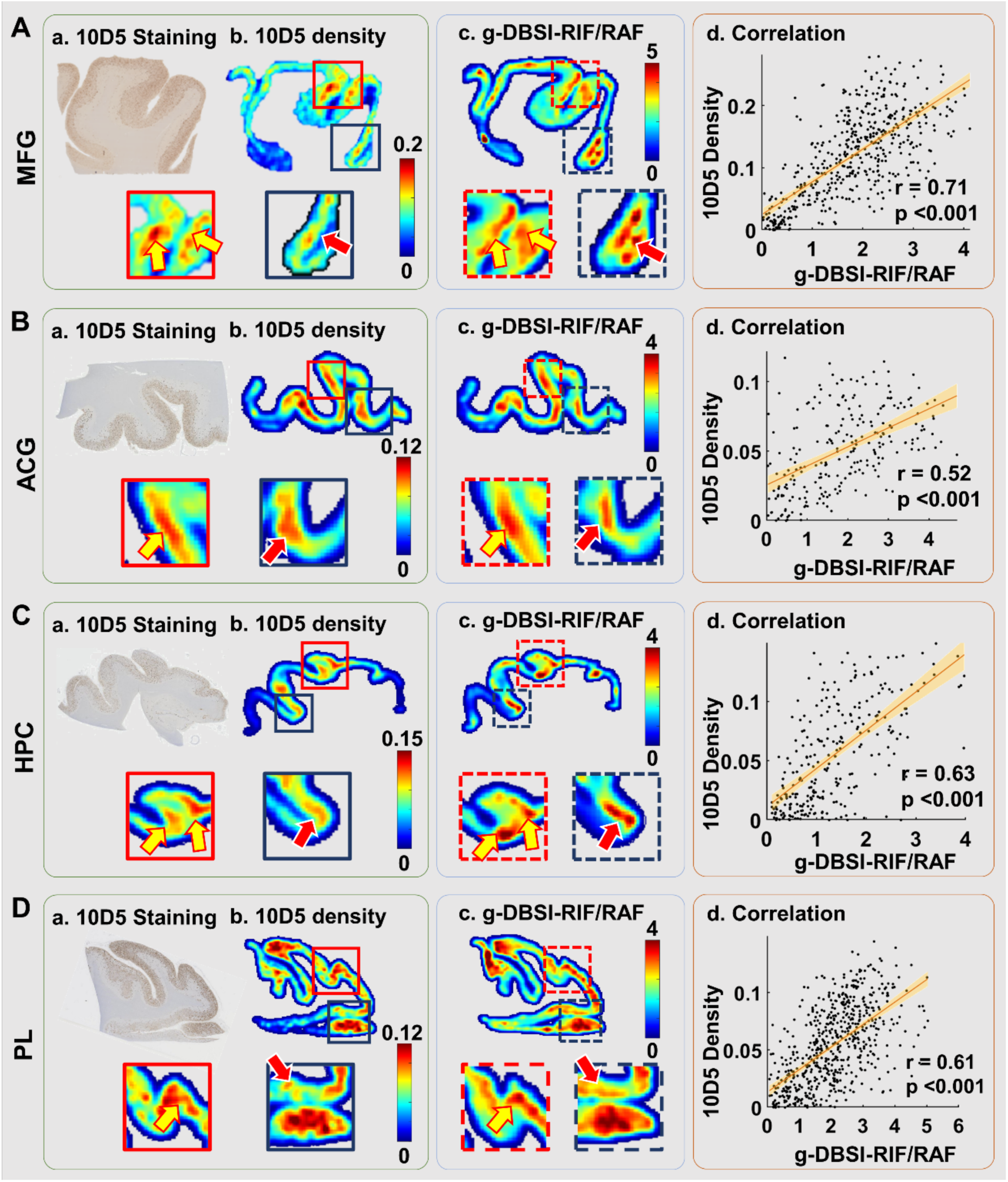
*Ex situ* g-DBSI RIF/RAF spatially correlates with amyloid-beta deposition across cortical brain regions in postmortem brain tissue blocks. Representative results from four brain regions: middle frontal gyrus (MFG, **A**), anterior cingulate gyrus (ACG, **B**), hippocampus (HPC, **C**), and parietal lobe (PL, **D**). For each region, (a) 10D5 IHC staining of the tissue section; (b) quantitative amyloid-beta density map derived from 10D5 staining; (c) co-registered g-DBSI-derived RIF/RAF map from the same tissue block; (d) region-wise scatter plot of 10D5 amyloid-beta density versus g-DBSI RIF/RAF, with the orange line indicating the least-squares linear regression fit and shaded band representing the 95% confidence interval. Red and blue dashed inset boxes in panels (a–c) denote representative subregions shown at higher magnification below each whole-section image. Yellow and red arrows indicate areas of spatially concordant elevated signal between 10D5 amyloid-beta density and RIF/RAF maps. Color bars indicate relative 10D5 amyloid-β density (b) and RIF/RAF values (c). Pearson correlation coefficients (r) and corresponding p-values are reported for each region (d; all *p* < 0.001).

To further characterize the spatial relationship between g-DBSI-derived RIF and 10D5 density, the individual RIF maps were examined for the same brain regions. We found the significant spatial correlations between g-DBSI-RIF and 10D5 density (**Fig. S3A-D**). The averaged correlation coefficient across all postmortem brain tissue blocks included in this analysis is 0.48 (SD = 0.11 (Mean ± SD), which is significantly lower than that between RIF/RAF and 10D5 density (*p* < 0.05).

### *In vivo* cortical microstructural g-DBSI measures correlate with PET amyloid burden differently in low and high amyloid deposition individuals

In our *in vivo* human cohort analysis, maps of cortical Pittsburgh compound B (PIB) PET standardized uptake value ratio (SUVR), a quantitative measure of amyloid-beta burden in humans (*22*), were co-registered with g-DBSI RIF/RAF maps from the same participants. To investigate whether cortical amyloid-beta burden relates to g-DBSI metrics *in vivo*, 112 participants from the Knight ADRC cohort were recruited to undergo PIB PET and g-DBSI MRI scans. Centiloid (CL) values were generated from PIB PET for this cohort. In line with recent CL recommendations(*23–25*), a cutoff of 12 CL was selected as an “early abnormality” threshold to capture emerging A*β* pathology. Prior PET and CSF studies have shown that a CL value of approximately 12-CL maximizes agreement with abnormal CSF Aβ42, consistent with the onset of early amyloid pathology (*23–25*). This lower threshold aligns with neuropathology-anchored work suggesting that values below about 10-12 CL indicate absence of amyloid pathology, whereas values at or above this range mark the transition from no to subtle amyloid pathology.

Participants were stratified into two groups: CL <12 (n=78) and CL >12 (n=34). FS segmentation was performed on the MPRAGE using version 6.0 (http://surfer.nmr.mgh.harvard.edu/). The whole cortical region was segmented, and the g-DBSI metrics were averaged within cortical regions. The correlations between g-DBSI metrics and PET CL were generated in two groups. Globally, amyloid burden quantified in CL showed differential associations with g-DBSI metrics below versus above the prespecified 12-CL threshold. Among individuals with higher amyloid burden (CL > 12, mulberry), greater CL values correlated with higher RIF/RAF (r = 0.35, p = 0.04; **Fig. 5Aa**) and lower RAF (r = −0.42, p = 0.02; **Fig. 5Ab**), while relationships with HAF, RIF, HIF, and PF are weak and did not reach statistical significance (**Fig. 5Ac-f**). In contrast, in participants with low amyloid (CL < 12, green), higher CL values were significantly associated with higher RAF (r = 0.33, p < 0.01; **Fig. 5Ab**), higher RIF (r = 0.23, p = 0.05; **Fig. 5Ad**), and lower HIF (r = −0.37, p < 0.01; **Fig. 5Ae**). Region-wise partial correlation analyses revealed that FDR-corrected associations between g-DBSI indices and amyloid CL were present exclusively in the amyloid-negative group (CL < 12) and absent in the amyloid-positive group (CL > 12, **Fig. 5B**). Any regions surviving FDR-correction are shown on Desikan-Killiany cortical surface maps in **Fig. S4**. In the low-amyloid group, RAF and HIF both showed positive correlations with CL, focused on default-mode network regions, particularly the posterior cingulate, precuneus, superior frontal, and rostral middle frontal cortex, as well as the superior temporal and auditory association areas. RIF showed FDR-corrected positive associations restricted to frontal regions (pars orbitalis and rostral middle frontal), and HAF to posterior cingulate and precuneus. RIF/RAF and PF yielded no FDR-corrected regional associations in either group.

**Figure 5.**
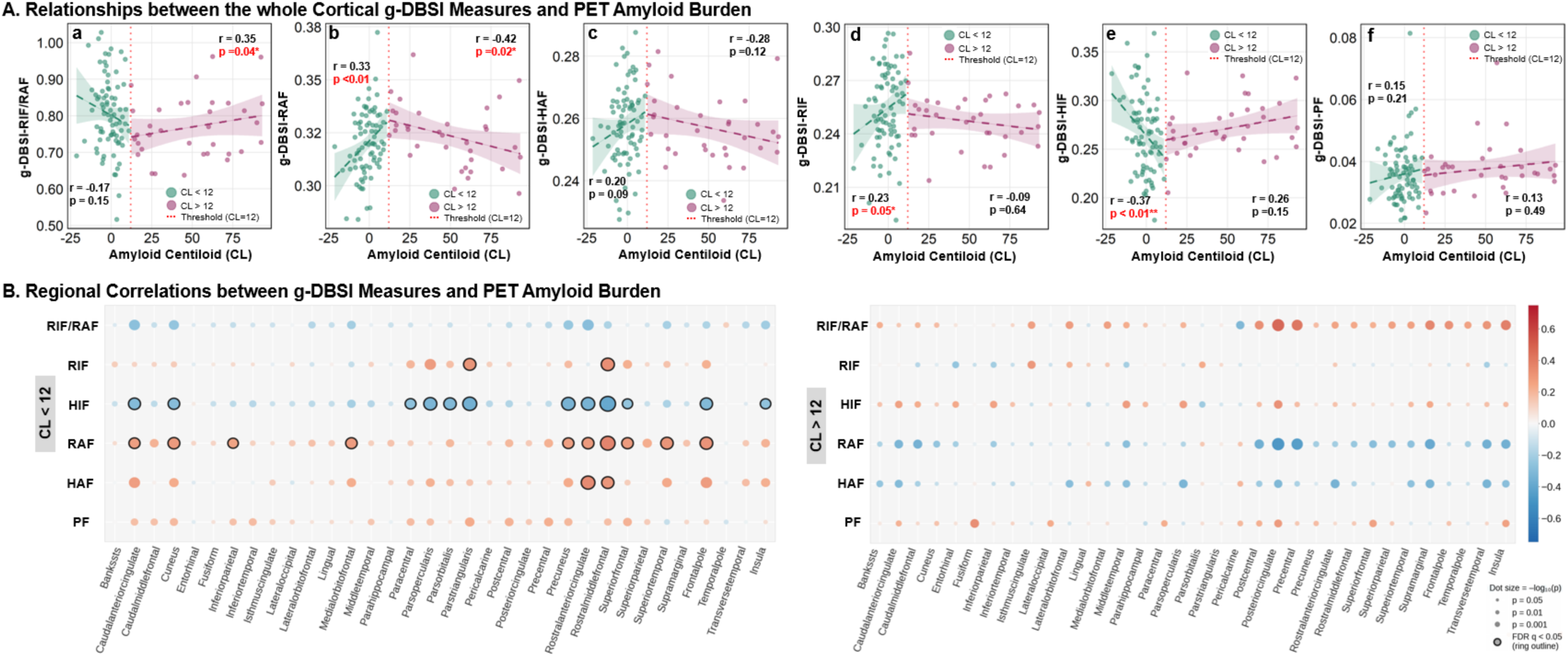
Amyloid burden-dependent relationships between cortical g-DBSI microstructural measures and PET amyloid deposition across low- and high-amyloid groups. (**A**) Scatter plots illustrate age- and sex-adjusted partial correlations between whole-cortex g-DBSI indices and amyloid burden (Centiloid, CL) quantified by PIB-PET, stratified by low- (CL < 12, green) and high-amyloid (CL > 12, pink) groups. Panels demonstrate RIF/RAF neuroinflammation index (a), restricted anisotropic fraction (RAF; b), hindered anisotropic fraction (HAF; c), restricted isotropic fraction (RIF; d), hindered isotropic fraction (HIF; e), and perfusion fraction (PF; f). Dashed lines indicate least-squares linear regression fits within each group; vertical red dotted line indicates the CL = 12 threshold. Pearson partial correlation coefficients (r) and p-values are shown for each group; *p < 0.05, **p < 0.01. (**B**) Bubble plots show region-wise age- and sex-adjusted partial correlations between individual g-DBSI indices and CL across cortical ROIs, separately for the low- (CL < 12, left) and high-amyloid (CL > 12, right) groups. Color encodes the direction and magnitude of the correlation (red: positive; blue: negative); dot size reflects -log10(p). Dots with ring outlines indicate regions surviving FDR correction (q < 0.05).

To assess the robustness of the primary findings, we conducted a sensitivity analysis using a CL cutoff of 20, which has been used by most studies to define the amyloid positivity (*26, 27*).

Participants were stratified into amyloid-negative (CL<20, n=85) and amyloid-positive (CL>20, n=27) groups. In the CL<20 group, RAF showed a significant positive correlation with CL (**Fig. S5B**, r = 0.40, *p* < 0.001), stronger than that observed in the CL<12 low-amyloid group (r = 0.33, *p* < 0.01). RIF/RAF showed a significant negative correlation with CL (**Fig. S5A**, r = −0.22, *p* = 0.05), and HAF showed a significant positive correlation with CL (**Fig. S5C**, r = 0.29, *p* < 0.01), neither of the other g-DBSI indices (RIF, HIF, and PF) reached significance in the CL<12 analysis (**Fig. S5D-F**). In the CL>20 group, no significant correlations were observed between g-DBSI indices and CL, though directional trends were consistent with the primary analysis.

The relationships between DTI-derived mean diffusivity (MD), a DTI index used to investigate cortical microstructural alterations in AD (*28, 29*), and amyloid CL were also explored in this study. Using the CL=12 threshold, DTI-MD showed a significant negative correlation with CL in the low-amyloid group (CL<12: r = −0.26, *p* = 0.02) but not in the high-amyloid group (CL>12: r = 0.15, *p* = 0.34) (**Fig. S6A**). Sensitivity analysis using CL=20 yielded consistent directional results that DTI-MD remained significantly negatively correlated with CL in the CL<20 group (r = −0.25, *p* = 0.02) but not in the CL>20 group (r = 0.12, *p* = 0.58) (**Fig. S6B**). Negative correlations between MD and CL were spatially distributed across frontal, parietal, and cingulate cortices (**Fig. S6C**).

### CSF proteomic signatures associated with the g-DBSI neuroinflammation index

To investigate the molecular correlates of g-DBSI-derived neuroinflammation, we explored 55 Knight ADRC participants (22 females, 23 males; mean age 68.9 ± 9.5 years; mean education 16.3 ± 2.1 years; 32.7% APOE-ε4 carriers) who underwent g-DBSI scans and CSF proteomic profiling using the SomaScan 7k platform. Among those, 45 participants were negative for amyloid deposition and had normal cognition. Eight participants were positive for amyloid deposition and had normal cognition, and two were diagnosed with AD dementia and were positive for amyloid deposition. Linear regression analysis was performed to assess the association between CSF proteomics and g-DBSI RIF/RAF, adjusting for age and sex. The volcano plot summarizes the strength and direction of these associations (**Fig. 6A**). In total, 200 proteins were significantly positively associated with g-DBSI RIF/RAF, whereas 278 proteins were significantly negatively associated (unadjusted *p* < 0.05).

**Figure 6:**
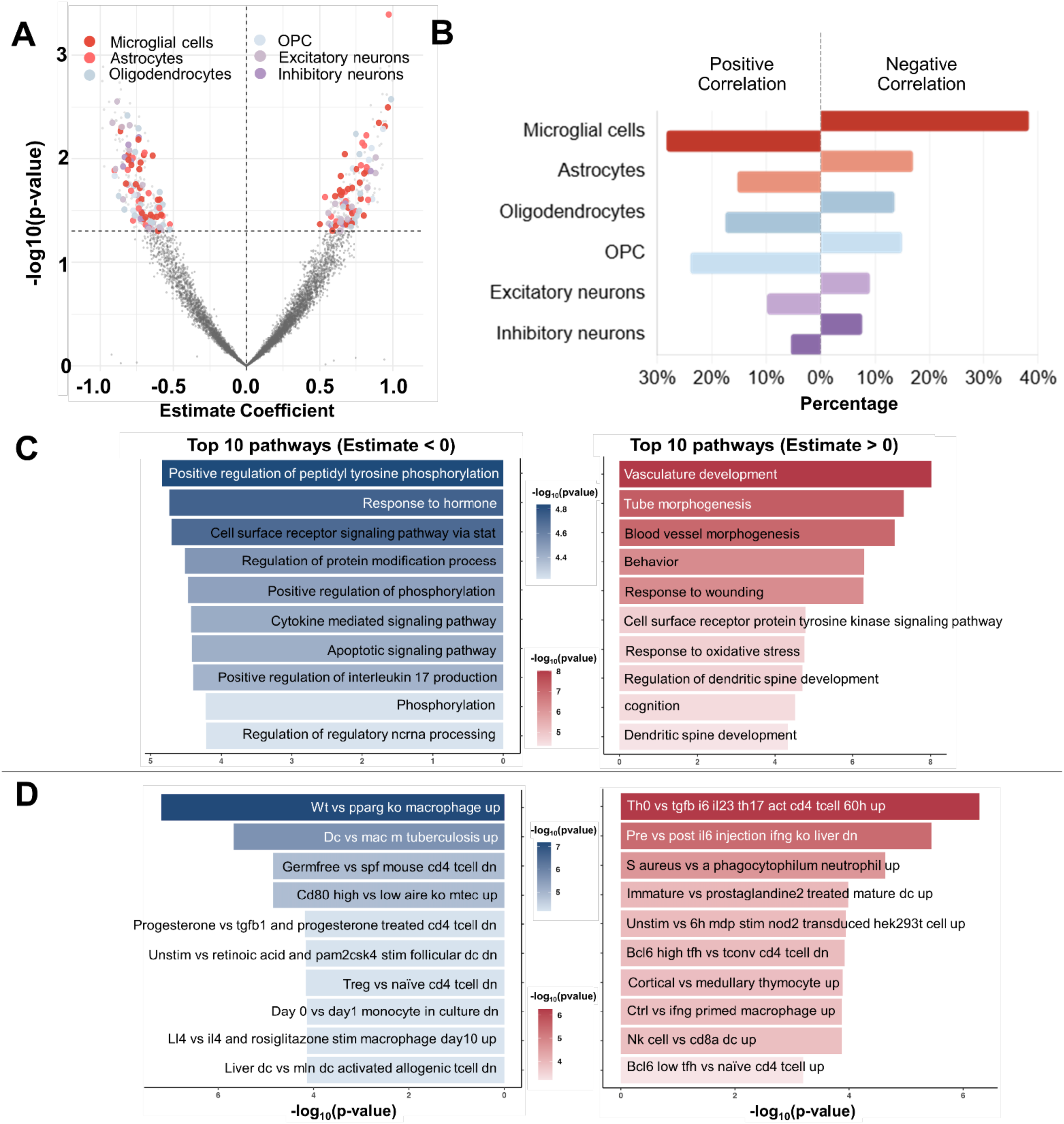
CSF proteomic signatures associated with the g-DBSI-derived RIF/RAF. **(A)** Volcano plot showing associations between CSF protein levels (SomaScan 7k) and whole-cortex g-DBSI RIF/RAF. Each point represents one protein; the x-axis shows the regression coefficient (estimate) and the y-axis shows −log10(p-value). Dashed horizontal and vertical lines indicate the nominal significance threshold (*p* = 0.05) and zero effect, respectively. Colored points represent proteins passing the nominal significance threshold, annotated by cell-type expression: microglial cells (dark red), astrocytes (salmon), oligodendrocytes (blue), oligodendrocyte precursor cells (OPC, light blue), excitatory neurons (light purple), and inhibitory neurons (purple). (**B**) Cell-type composition of RIF/RAF-associated proteins, separately for positively (left) and negatively (right) associated proteins, based on reference single-cell expression data. Bar lengths reflect the proportion of proteins assigned to each major brain cell class. (**C**) Top 10 enriched Gene Ontology (GO) biological processes among proteins negatively (left, blue) and positively (right, red) associated with RIF/RAF, ranked by -log10(adjusted p-value). Color intensity reflects enrichment significance. (**D**) Top 10 immune and perturbation signatures enriched among proteins negatively (left, blue) and positively (right, red) associated with RIF/RAF, highlighting transcriptional programs related to macrophage/microglial activation, T-cell differentiation, and cytokine signaling, ranked by -log10(adjusted p-value).

Cell-type annotation of RIF/RAF-associated proteins using the Human Protein Atlas (HPA), a publicly available dataset (*30*), revealed an over-representation of glial markers among the positive associations (**Fig. 6B**). Proteins with higher levels at higher RIF/RAF values were enriched for microglial and astrocytic signatures, with smaller contributions from oligodendrocyte-lineage markers. In contrast, proteins negatively associated with RIF/RAF showed a more mixed cellular profile, including oligodendrocyte precursor cells and neuronal markers. These distributions suggest that elevated RIF/RAF reflects a systemic proteomic footprint consistent with activated astrocytes and microglia.

Pathway enrichment analysis further distinguished positive and negative RIF/RAF associations (**Fig. 6C-D**). Top-enriched Gene Ontology biological processes (GO BP) among proteins negatively associated with RIF/RAF included pathways related to cytokine-mediated signaling, cell-surface receptor signaling via STAT, and apoptotic and interleukin-17-related processes, consistent with broad inflammatory activation. In contrast, positively associated proteins highlighted vascular development, tube and blood-vessel morphogenesis, responses to wounding and oxidative stress, and regulation of dendritic spine development and cognition (**Fig. 6C**). To further characterize the immune microenvironment, we evaluated specific immunologic signatures. From the immunologic signature gene sets, we found that lower RIF/RAF aligned with transcriptional programs induced by strong macrophage, whereas higher RIF/RAF tracked with more regulated T-cell and innate immune states (**Fig. 6D**). Collectively, these proteomic findings support the interpretation that g-DBSI RIF/RAF captures a glia-centered inflammatory phenotype that is embedded within broader immune and vascular signaling networks, while also emphasizing the need for replication in larger cohorts given the current study’s limited power.

## Discussion

This study integrates *ex situ* and *in vivo* dMRI, quantitative histopathology, and CSF proteomics to establish the biological basis of g-DBSI metrics in human AD brains. We demonstrate that g-DBSI separates microstructural compartments, cellularity (RIF), and neurofilaments (RAF), which correspond to their histological counterparts across seven cortical and subcortical regions. The neuroinflammation index RIF/RAF aligns with astrocytic and microglial pathology as well as glia-enriched CSF proteomic signatures. Together, these findings provide evidence that g-DBSI can noninvasively map inflammation-related tissue changes while preserving sensitivity to concurrent neurofilament alterations in both WM and GM.

This study provides a direct biological grounding for g-DBSI metrics by linking them to their corresponding histological structures. The RIF reflects water restricted within cellular structures with very low apparent diffusion coefficients (0 - 0.4 µm²/ms for *ex situ* tissue and 0-0.8 for *in vivo* brains), a signature consistent with densely packed inflammatory cells, activated microglia, and reactive astrocytes. Across all sampled regions, RIF correlated significantly with H&E-derived cellularity, confirming that this metric captures total cellular burden in both white and gray matter. Conversely, the RAF tracked neurofilament density (2F11), reflecting axonal tracts in white matter and dendritic arbors in gray matter. Crucially, while conventional diffusion MRI is largely restricted to assessing white matter tracts, these correlations demonstrate that g-DBSI is also highly sensitive to gray matter tissue changes related to neurofilament, accurately capturing the integrity of dendritic arbors, unmyelinated axons, and neuronal cell bodies in the cortex. These complementary mappings establish that g-DBSI decomposes the diffusion signal into biologically interpretable compartments rather than composite, tissue-nonspecific indices.

To enhance sensitivity to an inflammatory cell-focused proxy for neuroinflammatory remodeling, a g-DBSI index was defined as the ratio of RIF to RAF. This ratio reflects the balance between inflammatory cellularity-related restricted diffusion and organized neurofilament-associated diffusion and was used consistently in all subsequent *ex situ* and *in vivo* analyses. RIF/RAF colocalized with the summed GFAP and IBA-1 densities across all sampled regions in both WM and GM, indicating its sensitivity to spatial variations in astrocytic and microglial activation. During neuroinflammatory processes, microglial cells and astrocytes undergo morphological changes, including soma enlargement, process retraction or hypertrophy, and proliferation, which increase restricted diffusion while simultaneously displacing organized neural structures, thereby amplifying the RIF/RAF signal. This ratio-based design makes RIF/RAF more sensitive to early or subtle neuroinflammatory changes than either absolute RIF or RAF alone.

The spatial coupling between RIF/RAF and amyloid pathology observed in ex situ tissue blocks from donors with established, high-burden AD (Thal Stage 3-5) is mechanistically consistent with the inflammatory biology of advanced peri-plaque remodeling. In this phase, reactive microglia and astrocytes undergo soma enlargement, process retraction or hypertrophy, and net proliferation in direct response to dense amyloid deposits, thereby increasing water restriction within inflammatory cell bodies (as captured by RIF) while simultaneously displacing and disrupting the coherent neurofilament and dendritic architecture that underlie RAF. Together, these changes amplify the RIF/RAF ratio in amyloid-rich areas, as captured in **Figure 4**.

The superiority of RIF/RAF over RIF alone in capturing the spatial distribution of amyloid-related inflammation (**Fig. S3**) highlights a key advantage of the ratio-based g-DBSI marker. While RIF reflects total cellular burden, it cannot distinguish peri-plaque glial proliferation from residual cellularity in AD. RAF normalization suppresses contributions from tissue areas that retain organized neurofilament architecture, thereby sharpening the specificity of the inflammatory signal to regions actively undergoing glial-mediated remodeling. Moreover, the RIF/RAF ratio could mitigate the confounding effects of free water and vasogenic edema, which typically occur in the later stages of AD. In amyloid-burdened tissue, extracellular free water, which is captured primarily in the g-DBSI-derived HIF, can partially dilute or obscure RIF in absolute terms, as increased diffusivity from edema shifts the signal away from the restricted isotropic spectrum. Because edema affects both the RIF and the broader isotropic diffusion environment, normalization to RAF could make this marker largely insensitive to isotropic free-water content, and potentially provide a self-normalizing index that is more robust to edema-related confounds than RIF alone. This is particularly relevant in AD, where cerebrovascular dysfunction, blood-brain barrier leakage, and amyloid angiopathy can introduce regional free-water variation unrelated to glial cellularity per se (*31–34*). The lower variance of RIF/RAF across brain regions (0.11 vs. 0.19 for RIF) further demonstrates that the ratio provides a more regionally stable index to reflect inflammatory cell density. This regional stability is especially important in AD, where the balance between inflammatory remodeling and preserved neural infrastructure differs markedly across hippocampal and association cortices along Braak and Thal trajectories (*35–37*).

It is important to interpret the spatial correlation data in Figure 4 with two caveats in mind. First, the analysis necessarily reflects a cross-sectional and regionally averaged measure of co-occurrence between g-DBSI RIF/RAF and amyloid density; it does not resolve whether elevated RIF/RAF precedes, accompanies, or lags amyloid deposition in the temporal sequence of AD pathology, a question that can only be addressed longitudinally. Second, formalin fixation and the *ex situ* diffusion environment alter absolute diffusivity values compared to in vivo conditions, though the relative spatial patterns of restricted vs. hindered diffusion are expected to be preserved.

In our *in vivo* study, our data demonstrated amyloid burden-dependent relationships between whole-cortical g-DBSI microstructural measures and PET amyloid CL: an early positive association between amyloid burden and neurofilament-related (RAF) and cellular (RIF) signals, followed by a later shift toward rising RIF/RAF and declining RAF and RIF. A large body of structural imaging work supports this framework. A study of cortical microstructural changes across the AD continuum in a large multicenter cohort found that cortical mean diffusivity, free water, and cortical thickness followed a biphasic trajectory, with early preclinical AD characterized by cortical thickening and reduced mean diffusivity, which is consistent with cellular proliferation, and later stages showing progressive diffusivity increases reflecting neurodegeneration (*38*). Consistent with this, other studies also demonstrated the non-monotonic relationship between AD pathology and dMRI measures of brain microstructure (*29, 39*). The present g-DBSI results extend these observations by decomposing cortical diffusion signals into biologically interpretable components, providing a mechanistically richer view of how inflammation- and degeneration-related microstructural changes evolve along this trajectory.

In the early phase of AD, when the tissue is largely intact, an increase in both RAF and RIF in low-amyloid individuals may reflect initial compensatory dendritic sprouting (*38, 40*), astrocyte process hypertrophy (*41, 42*), intermediate fibrillary astocytosis(*43*), and hyper-ramified microglial process extension (*44, 45*). In contrast, in the high-amyloid group, the simultaneous rise in RIF/RAF and fall in RAF suggests that progressive glial proliferation and soma enlargement are occurring at the expense of organized neurofilament architecture.

In this study, CSF proteomic profiling was explored to independently validate the biological specificity of RIF/RAF. Our findings provide the initial proteomic evidence directly linking this imaging index to a definable and mechanistically coherent neuroimmune state, substantially advancing the biological interpretability of g-DBSI-RIF/RAF as a translational biomarker. The enrichment of pathways such as "response to wounding," "response to oxidative stress," and "vasculature development" perfectly aligns with the physiological scenarios of the inflamed brain microenvironment in AD. In the CNS, vascular remodeling and cellular stress responses are necessary structural adaptations that facilitate permeability of the blood-brain barrier and subsequent infiltration of peripheral immune cells. This mechanism is consistent with and supported by our elevated g-DBSI-RIF/RAF in association with aggressive, tissue-infiltrating effector states, including Th17 activation and IFN-γ-primed macrophage responses. Critically, the concurrent suppression of cytokine-mediated signaling, JAK-STAT activation, apoptotic clearance, and regulatory macrophage programs indicates that this infiltrative state is not self-resolving but rather reflects a failure of immunoregulatory containment, in which peripheral immune cells persist and sustain the elevated g-DBSI signal over time. Proteins associated with dendritic spine development and cognition were also enriched at higher g-DBSI-RIF/RAF values, potentially linking neuroinflammation to synaptic damage. All those findings suggest g-DBSI-RIF/RAF as a translational biomarker capable of capturing the complex cellular alterations underlying neuroinflammatory processes in AD brains. Given the exploratory nature of this analysis and the absence of formal multiple-testing correction in the current sample, these findings should be validated by future larger studies.

Regarding the translational implications, g-DBSI is radiation-free, compatible with clinical scanners, and amenable to longitudinal sampling, properties that position it for deployment in therapeutic trials where glial responses determine both efficacy and risk. RIF/RAF could potentially inform patient stratification and safety monitoring for anti-amyloid monoclonal antibody therapies, in which amyloid-related imaging abnormalities (ARIA) reflect neuroinflammatory and vascular sequelae that current conventional MRI metrics cannot mechanistically resolve. FF and CF provide complementary readouts of axonal integrity and cellular burden, together enabling mechanistic phenotyping across regions and disease stages. The observed molecular anchoring to CSF proteins and pathways further suggests a route toward integrated MRI–proteomic endpoints that link brain microstructure to systemic immune programs.

Key strengths of this study include (i) voxel-wise MRI-histology registration across multiple cortical and subcortical AD-vulnerable brain regions, (ii) orthogonal inflammatory cell markers (GFAP, IBA-1) alongside cellular (H&E), axonal (2F11), and amyloid-beta peptide (10D5) stains, and (iii) independent molecular anchoring via CSF proteomics with cell-type and pathway enrichment. Convergence across these modalities increases confidence in the biological specificity of g-DBSI metrics. However, several limitations warrant consideration. First, co-registration between MRI and histology is inherently challenging because fixation and sectioning introduce tissue deformation; although block-face–guided elastic registration and quality-control exclusion criteria were applied, residual misalignment cannot be fully eliminated. Second, the 6 µm thickness of histology sections versus the 1 mm MRI slice creates a partial-volume mismatch that may attenuate correlations; future work using serial-section aggregation or volumetric histology (e.g., CLARITY-based tissue clearing with light-sheet microscopy) could provide thickness-matched validation. Third, the *ex situ* validation cohort was small (six donors, 42 blocks), limiting generalizability across the full range of AD severity and co-pathologies. Fourth, the CSF proteomic analyses used unadjusted significance thresholds and a modest sample size (n = 55), necessitating replication in larger, independent cohorts with appropriate multiple-testing correction. Fifth, the cross-sectional design precludes assessment of whether RIF/RAF changes precede, accompany, or follow amyloid accumulation and cognitive decline; prospective longitudinal studies are needed to establish the temporal dynamics of g-DBSI metrics along the AD continuum. Finally, the overcomplete diffusion dictionary in g-DBSI, while more flexible than fixed-compartment models, still relies on prior assumptions embedded in the tensor bases and in the Monte Carlo simulations used to construct it; sensitivity analyses that vary dictionary parameters would further establish robustness.

Future work should standardize g-DBSI acquisition and processing across scanner platforms, extend validation to prospective multisite cohorts, further calibrate *ex situ*-to-*in vivo* parameter harmonization, and test whether RIF/RAF predicts clinical outcomes and treatment-emergent adverse events such as ARIA. Integration of g-DBSI with tau PET and longitudinal fluid biomarkers will clarify how neuroinflammation interacts with the broader AD pathological cascade. This translational framework supports near-term deployment of g-DBSI for patient stratification and longitudinal monitoring and sets the stage for integrative MRI–proteomic readouts that bring cell-type specificity to routine neuroimaging.

## Conclusion

In this cross-sectional, multimodal study, we provide converging evidence from histopathology, MRI, PET, and CSF proteomics that g-DBSI yields biologically specific readouts of neuroinflammation and neurodegeneration in AD. Co-registration of g-DBSI maps with quantitative immunohistochemistry across seven important AD-vulnerable brain regions demonstrated that the RIF derived from g-DBSI robustly reflects total cellularity, while the RAF tracks neurofilament integrity in both white and gray matter, establishing that g-DBSI decomposes the diffusion signal into histologically grounded compartments. The neuroinflammation index, defined as the ratio of RIF to RAF, colocalized with astrocytic and microglial densities in peri-plaque tissue and scaled with cortical amyloid burden *in vivo*, revealing a potential biphasic relationship that suggests a transition from early compensatory cellular responses to progressive neurodegeneration at higher amyloid loads. Independent CSF proteomic profiling further anchored the neuroinflammation index to glia-enriched immune and vascular signaling pathways, providing the first molecular evidence that this imaging metric captures a mechanistically coherent neuroinflammatory state. Although replication in larger, longitudinal, multisite cohorts is needed to establish generalizability and temporal dynamics, the radiation-free, clinically compatible nature of g-DBSI positions it as a promising translational biomarker for patient stratification, longitudinal monitoring of immune responses, and risk monitoring of treatment-related adverse events such as amyloid-related imaging abnormalities in current and future AD clinical trials.

## Materials and Methods

### Study design

This was a cross-sectional, multimodal validation study designed to establish biological correlations among postmortem tissue pathology, *ex situ* and *in vivo* dMRI metrics, *in vivo* PiB PET imaging, and proteomic signatures of neuroinflammation in AD. The study comprised three arms: (1) *ex situ* MRI-histology validation in postmortem brain tissue blocks, (2) *in vivo* g-DBSI analysis across AD stages with amyloid PET co-registration, and (3) CSF proteomics profiling underlying g-DBSI-derived neuroinflammation index. The postmortem AD brains were donated by participants of the Knight ADRC. After formalin fixation, small samples (blocks) from those postmortem brain specimens underwent g-DBSI MRI scans and, subsequently, histologic processing and staining with H&E for cellularity, 2F11 IHC for neurofilaments, GFAP IHC for astrocytes, IBA-1 IHC for microglia, and 10D5 IHC for amyloid-beta deposits. Stained six-micron slide-mounted tissue sections were digitized and spatially registered to *in vivo* dMRI to enable region-of-interest (ROI)-level correlations. The relationship between the g-DBSI metrics and amyloid PET SUVR was investigated in living human participants. CSF proteomics has been used to identify biological pathways underlying the g-DBSI-derived neuroinflammation index.

All procedures were approved by the Washington University Institutional Review Board through the Human Research Protections Office and performed in accordance with the Declaration of Helsinki. Written informed consent was obtained from all participants for the use of *ex situ* specimens and in vivo data.

### Postmortem brains tissue samples

Thirty-five formalin-fixed postmortem brain tissue blocks from six Knight ADRC participants (characteristics summarized in **Table S1**), representing seven different brain regions, were included in this study to validate the imaging findings of g-DBSI. Within 24 hours of expiration, the left hemisphere of each brain was thoroughly fixed in 10% neutral-buffered formalin (*46*) and samples, each approximately 15 × 20 × 6 mm, were excised from seven regions, including 1) Middle frontal gyrus; 2) Anterior cingulate gyrus; 3) Striatum with nucleus accumbens and olfactory cortex; 4) Hippocampus with parahippocampal, fusiform, inferior temporal gyri; 5) Posterior cingulate gyrus and precuneus; 6) Parietal lobe (angular gyrus); 7) Occipital lobe (calcarine sulcus and parastriate cortex) (**Fig. S1**).

### Living Study participants

All living participants recruited from the Knight ADRC Memory and Aging Project cohort at Washington University School of Medicine (St Louis, MO, USA) (1) were in the age range of 50 to 100 years; 2) had undergone a g-DBSI dMRI scan; and 3) had undergone PET amyloid within one year of the g-DBSI acquisition. The participants’ flow diagram is shown in **Fig. S7**. The demographics of those participants are summarized in **Table 1**. The demographics of a subset of participants with CSF proteomics collected within one year of the g-DBSI acquisition were summarized in **Table S2**.

**Table 1.** Demographics and Clinical Characteristics of Participants.

| Variable | CL < 10 | CL > 10 | P-value |
| --- | --- | --- | --- |
| N | 77 | 35 | - |
| CDR=0/CDR>0 | 77/0 | 30/5 | - |
| Age | 67.9 ± 8.1 | 73.1 ± 7.9 | **0.002** |
| Female, n (%) | 39 (50.0%) | 19 (54.3%) | 0.827 |
| MMSE | 29.4 ± 1.2 | 28.8 ± 1.7 | **0.030** |
| CDR Sum of Boxes | 0.03 ± 0.23 | 0.49 ± 1.18 | **0.001** |
| PACC | 0.304 ± 0.603 | -0.166 ± 0.642 | **<0.001** |
| APOE ε4+, n (%) | 21 (26.9%) | 21 (60.0%) | **0.002** |
| Amyloid Centiloid | -1.50 ± 6.11 | 48.17 ± 26.85 | **<0.001** |

Note: Values are mean ± SD for continuous variables and n (%) for categorical variables. P-values are from independent t-tests for continuous variables and chi-square or Fisher's exact test for categorical variables. Bold p-values indicate statistical significance ( $p < 0.05$ ).

### g-DBSI Modeling

g-DBSI relies on endogenous signals from water diffusion to probe microstructural changes associated with neuroinflammation and neurodegeneration in AD. Specifically, g-DBSI employs dMRI sequences with a multi-directional, multi-weighting diffusion protocol to measure signal decay due to water diffusion. The dMRI signal from a single imaging voxel (orange box in **Fig. 7A**) can be organized into a 1D vector according to the diffusion protocol. We denote the dMRI signal by an N×1 vector d (brown curve in **Fig. 7B**). g-DBSI develops an extensive, overcomplete diffusion dictionary (denoted by a matrix *DD*, row number, *L* = 1600) to contain a large number of coherent atoms of dMRI signals from a comprehensive list of the cerebral anatomical and pathological components (anisotropic white and gray matter tissues, inter-neuronal, intra-neuronal, intra-cellular, extra-cellular water component, and perfusion, etc.).

**Figure 7:**
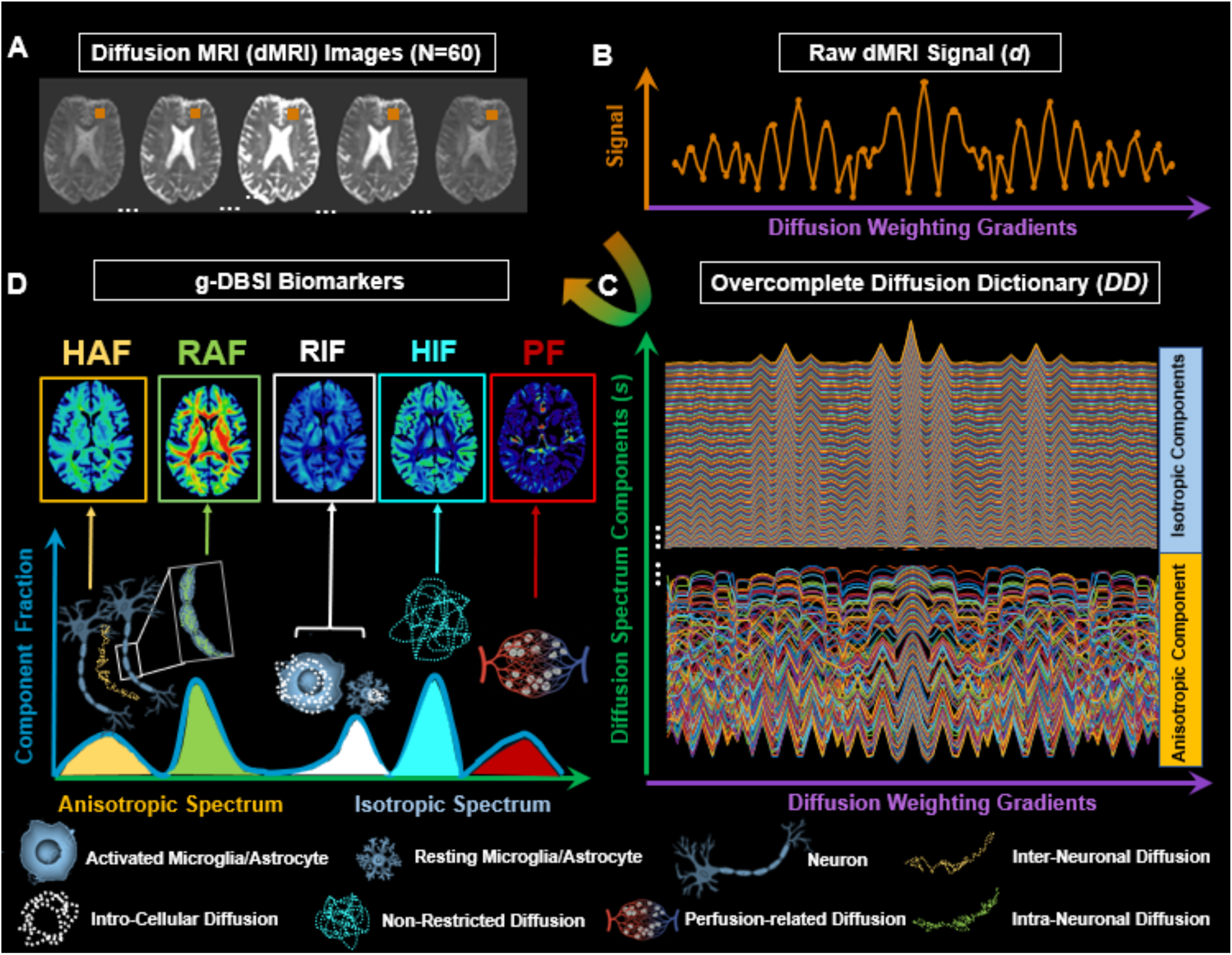
Schematic illustration of generalized diffusion basis spectrum imaging (g-DBSI) modeling framework for deriving diffusion-based microstructural biomarkers. (**A**) Multiple diffusion MRI (dMRI) images (N=60) are acquired using an FDA-approved dMRI sequence across varying diffusion-weighting gradients and b-values. A representative voxel (orange box) is selected to illustrate the signal modeling procedure. (**B**) The raw dMRI signal (*d*) within each voxel varies across diffusion-weighting gradients, reflecting contributions from multiple underlying tissue compartments with distinct diffusion properties. (**C**) g-DBSI decomposes the measured voxel signal by fitting it to an overcomplete diffusion dictionary (DD) comprising both anisotropic (fiber-like, bottom) and isotropic (cellular and fluid-like, top) spectrum components spanning a wide range of diffusivities and orientations. This dictionary-based decomposition enables the simultaneous estimation of low-to-high anisotropy and low-to-high isotropic-diffusivity compartments within each voxel. (**D**) The resulting diffusion spectrum decomposition yields five quantitative g-DBSI biomarker maps: hindered anisotropic fraction (HAF, reflecting inter-neuronal diffusion), restricted anisotropic fraction (RAF, reflecting intra-neuronal/axonal diffusion), restricted isotropic fraction (RIF, reflecting intra-cellular diffusion), hindered isotropic fraction (HIF, reflecting extra-cellular non-restricted diffusion), and perfusion fraction (PF, reflecting perfusion-related diffusion). These metrics provide *in vivo* characterization of biologically meaningful microstructural features, such as neurofilaments/axonal integrity, neuroinflammation, and cerebrovascular perfusion at the voxel level.

Each row of the *DD* represents an atom’s dMRI signal from a specific microstructural component type, with a particular orientation and diffusion pattern. Classic diffusion tensor theory (*47*) and Monte Carlo (MC) simulation (*48–50*) are used to compute each atom of the DD using the same diffusion protocol (DD column number = *N*). The details of MCS for water diffusion in cells are described in the supplement method section.

With the introduction of DD, the raw dMRI signal d (**Fig. 7B**) can be described as the weighted linear summation of the overcomplete atoms in DD (Eq. 1), where s is the L×1 positive weighting vector, e is an L×1 noise vector. The DD (L×N) formulation converts the high-dimensional fitting problem typically addressed by conventional dMRI methods into a large, ill-posed linear matrix inversion problem.

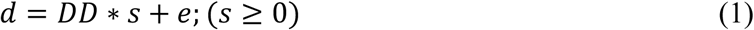

Due to the redundant property of the overcomplete dictionary, the weighted summation described in **Eq. 1** has a flexible and sparse representation, which can be utilized by the CS technique to effectively compute an accurate microstructural spectrum (the weighting factor, *s* in Eq. 1, blue spectrum in **Fig. 7D**) from a small number of clinical dMRI images (*51, 52*). The flexibility of building the overcomplete DD also increases the “illness” of the ill-posed CS problem. Fortunately, the microstructural spectrum is a nonnegative signal that can serve as additional strong prior knowledge for CS computation, helping avoid overfitting. Specifically, the non-negative CS formulation is presented in **Eq. 2**.

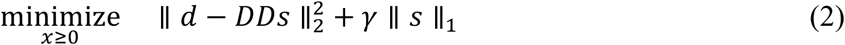

where the first term is the data consistency term in *L_2_* norm space, and the second term is the sparsity constraint in *L_1_* norm space. *γ* is a positive constant controlling the degree of sparsity of ***s***. Note that *γ* It depends on several factors, such as signal sparsity and the level of measurement noise in the signal. In this work, we will use the adaptive lasso method (*53*) to find *γ*, and solve **Eq. 2**. Similar methods have been used successfully in other fields such as economics, hyperspectral unmixing, face recognition, etc.

In g-DBSI, the entire microstructural components are grouped into five categories based on their anatomical and diffusion features: (1) inter-neuronal diffusion compartments with high anisotropy (yellow in **Fig.7D**); (2) intra-neuronal diffusion compartments with low anisotropy (green in **Fig.7D**); (3) restricted isotropic diffusion compartments with low diffusivity (white in **Fig.7D**); (4) non-restricted isotropic diffusion compartments with high diffusivity (cyan in **Fig.7D**); and (5) perfusion compartments that are sensitive to fast pseudo-diffusion effects (*54*) (red in **Fig.7D**).

### Monte Carlo simulation of intracellular water diffusion

To establish the biophysical link between cell-type-specific morphology and the diffusion MRI signal components modeled in g-DBSI, Monte Carlo simulations (MCS) of restricted intracellular water diffusion were performed for the three principal cell types relevant to AD neuroinflammation: astrocytes, microglia, and neurons. Realistic three-dimensional cellular geometries were obtained from GFP confocal image stacks available from NeuroMorpho.org, a publicly accessible and comprehensive repository of digitally reconstructed neurons, astrocytes, and microglia. Individual cell morphologies, including soma and processes, were semi-automatically segmented (**Fig. S8A**, top panels). In our MSC, 57 astrocytes, 970 microglia, and 148 neurons were included. Each segmented morphology was then reconstructed into a watertight 3D surface mesh (**Fig. S8A**, bottom left) and imported into the MCS framework (**Fig. S8A**, bottom right). Within each reconstructed cellular volume, restricted diffusion was modeled by initializing thousands of random walkers within the intracellular space and propagating them via stepwise Brownian motion under reflective boundary conditions at the cell membrane (**Fig. S8B**). For every diffusion-encoding gradient direction and b-value in the same acquisition protocol as the *ex situ* MRI scan, the accumulated phase of each walker was computed and ensemble-averaged across all walkers to generate synthetic diffusion-weighted signals, from which apparent diffusion coefficient (ADC) distributions were derived at 20 °C, which is the room temperature in the scanner (**Fig. S8B-C**). Since cells in tissue are randomly oriented with respect to the scanner gradient frame, the population-averaged diffusion signal is expected to be isotropic. Consistent with this, ADC distributions along the x-, y-, and z-axes are shown as representative orthogonal directions. There are modest direction-dependent shifts that did not alter the mean ADC or the shape of the distribution for all three cell types (**Fig. S8C**). The resulting ADC probability density distributions revealed cell-type-specific diffusion signatures: astrocytes exhibited a narrow ADC distribution centered at 0.236-0.240 µm²/ms, microglia at 0.356-0.368 µm²/ms, and neurons at 0.340-0.370 µm²/ms. The step number was set to 8,000, and the random seed count to 12,000 to ensure stable estimation of diffusion-weighted signals and derived metrics. These cell-type-dependent ADC distributions provided mechanistic priors for selective extraction of the restricted isotropic signal attributable to cellular components.

### Imaging Acquisition and Processing

#### Ex situ dMRI

Before the g-DBSI scan, the formalin-fixed tissue blocks were buffer-exchanged into phosphate-buffered saline (PBS) for 7 days at 4°C for rehydration purposes. The tissue blocks were embedded in 2% agar gel to stabilize them during the MRI scan and placed in a custom specimen holder. The prepared tissue blocks were scanned using a high-performance Helmholtz pair surface coil (*55*) on the Agilent/Varian DirectDrive 11.74-T (500-MHz) MRI system at the small animal MR facility at WashU. Acquisition parameters for g-DBSI are the following: TR = 1000ms, TE = 31.6ms, imaging resolution: 0.25×0.25×1mm^3^. To maintain consistency with the in vivo diffusion MRI protocol described below, *ex situ* dMRI was performed using a similar acquisition design, adjusted for the higher diffusion weighting required in postmortem tissue. The maximum b-value was 4500 s/mm², and diffusion gradients were applied in 28 directions across eight b-values (0, 100, 200, 500, 1000, 2000, 3000, and 4500 s/mm²). The acquisition time is approximately 9 hours to acquire sufficient SNR for each brain specimen. *Ex situ* dMRI data underwent signal drift correction (*56*), implemented in Python by modeling the global signal trend over time and normalizing accordingly to ensure temporal consistency. Following preprocessing, g-DBSI modeling was performed using an in-house MATLAB algorithm to decompose the diffusion signal into discrete isotropic and anisotropic components, enabling quantification of underlying microstructural features such as cellularity and axonal density. The resulting parametric maps were further used for cross-modal validation against histological measurements.

#### In vivo dMRI

The *in vivo* dMRI was acquired using multiple Siemens-built-in diffusion-vector schemes, including 6-, 10-, and 12-direction schemes. The 6-direction acquisition included b-values of 100 and 2000 s/mm², the 10-direction acquisition included b-values of 50, 500, and 1500 s/mm², and the 12-direction acquisition included b-values of 100 and 1000 s/mm². Combining all three sessions yielded 28 unique directions with 66 unique diffusion weightings. Each session included one non-diffusion-weighted volume (b = 0 s/mm²). The total acquisition time is approximately 9 minutes. The protocol was identical to that established for the Knight ADRC (*57*) and Dominantly Inherited Alzheimer Network (DIAN) studies (*58*). dMRI data were preprocessed using the DESIGNER2 pipeline (v2.0.13) run in a Singularity container, which performs denoising, Gibbs ringing removal, eddy-current and motion correction, and susceptibility distortion correction, followed by interpolation to an anatomically corrected (ANTs-based) space to generate an artifact-reduced 4D diffusion-weighted imaging volume. A brain mask was derived from the corrected b0 image using FSL’s BET with a fractional intensity threshold of 0.2 and robust center estimation. The preprocessed dMRI data and corresponding brain mask were then used as inputs for downstream g-DBSI modeling and statistical analysis.

#### Amyloid PET imaging

Two amyloid-PET tracers were used: ¹¹C-Pittsburgh Compound B (PIB) (*59*) and 18F-Florbetapir (FBP) (*60*). Within the XNAT platform on CNDA60, we have established a PET Universal Pipeline for generating standardized uptake value ratios (SUVRs) (*61*). SUVRs are analyzed using FreeSurfer regions of interest generated using the corresponding MRI MPRAGE, using a cerebellar cortex reference region. Data are generated with and without a partial volume correction implemented using a geometric transfer matrix approach (*62*). The global centiloid composite (CL) was calculated from the mean lateral orbitofrontal, medial orbitofrontal, precuneus, rostral middle frontal, superior frontal, superior temporal, and middle temporal regions (*63*).

#### Test-retest repeatability

To assess the repeatability of *ex situ* g-DBSI-derived metrics, five postmortem brain tissue blocks were randomly selected and scanned twice under identical acquisition conditions, including the same scanner hardware, pulse sequence parameters, and specimen positioning. and the resulting g-DBSI parametric maps were compared across time points. Quantitative repeatability was assessed voxel-wise using squared Pearson correlation coefficients (R²) between paired timepoint measurements as an index of spatial agreement and mean absolute deviation as an index of systematic measurement drift across repeated acquisitions. These metrics were computed separately for each g-DBSI-derived index to characterize repeatability. g-DBSI-derived parametric maps showed visually consistent spatial distributions of values between the two scan sessions. Voxel-wise comparison yielded R² values ranging from 0.85 to 0.97 and mean absolute deviations ranging from 0.014 to 0.029 across all g-DBSI metrics, indicating high spatial reproducibility under identical *ex situ* acquisition conditions.

### Quantitative Histology and MRI-histology Registration

After *ex situ* dMRI scans, specimens were returned to 10% formalin and processed into paraffin in the same orientation as in the MRI specimen holder. Sections (6 µm) were cut with a microtome from the bottom plane corresponding to the MRI bottom coronal plane, mounted on slides, and stained with hematoxylin & eosin (H&E), and immunohistochemistry for astrocytes (GFAP), microglia (IBA-1), neurofilament (2F11), and amyloid-beta peptide (10D5). Slide images were digitized at 20× magnification using a Leica Biosystems Aperio AT2 scanner.

High-resolution digital scans of stained tissue sections were analyzed using FIJI (ImageJ) for segmentation and quantification. Color deconvolution was applied to separate stain components, enabling selective isolation of target microstructure (nuclei, axonal fibers, microglia, and astrocytes) from the background. Segmented structures were thresholded and binarized, and particle analysis was conducted to quantify features of interest. For cellular analysis, individual nuclei were counted and normalized by area to generate two-dimensional number-density maps representing spatial distributions of cellularity. Similarly, axonal, microglial, and astrocytic densities were derived from the segmented stained images.

g-DBSI maps were first computed from the preprocessed *ex situ* diffusion MRI data. High-resolution histology images were then downsampled to match the dMRI resolution, and serial block-face photographs (taken of the exposed surface of the tissue block before each section was cut) were used as an intermediate reference to guide rigid and non-linear registration of each stained section back into the undistorted block geometry and into alignment with the corresponding dMRI slice. Registration quality was assessed using landmark error and tissue-mask overlap, The resulting co-registered voxel grids and white/gray matter masks provided a common space for predefined, region-of-interest analyses comparing g-DBSI metrics with histological measures across brain regions.

### Proteomics data collection and processing

Antemortem cerebrospinal fluid proteomics was performed using the same SomaLogic 7K SOMAscan aptamer-based platform and processing workflow described by Muhammad et al (*64*). CSF from our cohort (N=55 [Supplementary Table 2]) was collected by morning lumbar puncture after an overnight fast and stored at -80 °C until analysis. Protein abundance was quantified as relative fluorescence units (RFU) for the 7K SOMAscan panel, and SomaLogic’s standard pipeline was applied, including hybridization control-based intra-plate normalization, median-signal inter-plate normalization, and additional normalization to an external reference to reduce technical and biological variability. Aptamer- and sample-level quality control followed the published in-house pipeline: aptamers were excluded if calibration–median scale factor differences exceeded 0.5, median coefficient of variation was greater than 0.15, or log10 RFU values fell outside 1.5× the interquartile range in more than 85 % of samples; non-human targets were also removed. Samples were excluded when their log10 RFU profile was outside 1.5× the interquartile range for more than 85 % of aptamers, yielding a final dataset of 2,286 samples and 7,029 aptamers mapping to 6,163 unique proteins.

## Statistical Analysis

For the *ex situ* study, a 1.25 x 1.25 mm^2^ grid-box ROI across the brain blocks was implemented to evaluate the spatial correlations between the registered histology density maps and g-DBSI metrics. The number of grid boxes varied according to specimen size. For each ROI, mean histology density values and corresponding g-DBSI metrics were extracted. The spatial correlations between g-DBSI metrics and postmortem AD brain blocks were assessed using two-sided Pearson correlation at a 0.05 significance level for all cases.

To investigate the relationship between g-DBSI-derived brain microstructural metrics and amyloid burden, we performed correlation analyses stratified by PET amyloid CL thresholds. Participants were divided into two groups based on amyloid CL: CL < 12 and CL > 12. For each group, we calculated Pearson correlation coefficients between CL values and six g-DBSI metrics (RIF/RAF, RIF, HIF, RAF, HAF, and PF). Statistical significance was set at p < 0.05, with correlations reported as r values and corresponding p-values for each metric. Linear regression models with 95% confidence intervals were fitted separately for each amyloid group to visualize the divergent relationships. This stratified approach enabled detection of distinct relationships between amyloid pathology and brain microstructural changes that would be obscured in combined analyses, revealing critical threshold effects at the transition from early to later neurodegenerative processes. Cortical regions of interest were defined using the Desikan–Killiany cortical atlas implemented in FreeSurfer (*65*). Age and sex were adjusted as covariates for all analyses.

To identify proteins associated with g-DBSI-RIF/RAF in the cortical regions, differential protein abundance was first assessed using a linear regression model with age and sex as covariates. The lm function from the base stats package in R (v4.5.1) was used to fit the model to log10-transformed protein levels. The unadjusted p-values were reported. To annotate the cell-type specificity of candidate proteins (*66*), a publicly available dataset, the Human Protein Atlas (HPA) (*30*) was used. To assess cell-type specificity of SomaScan 7k targets, we leveraged human brain transcriptomic profiles from six major cell types (astrocytes, microglia, oligodendrocyte precursor cells, oligodendrocytes, excitatory neurons, and inhibitory neurons). For each cell type, we computed the mean expression of each gene, then summed these means across all six cell types to obtain the total expression per gene. We then derived cell-type expression proportions by dividing each cell type’s mean expression by the total expression for that gene. Genes were annotated as cell-type–specific when the highest cell-type proportion was at least 1.5-fold greater than the second-highest proportion; otherwise, genes were labeled as non-specific. SomaScan aptamers were mapped to their target genes and inherited the corresponding cell-type annotation, which was subsequently used for downstream cell-type enrichment summaries. Functional enrichment analysis was then performed separately for each group using the “clusterProfiler” function (*67*). We focused on pathway collections from the Immunologic signature gene sets and Gene Ontology Biological Process (GO BP) databases (*68, 69*). For each database and protein group, we reported the top 10 most significantly enriched pathways, or all pathways with an FDR-adjusted p-value < 0.05.

## Data Availability

The datasets used and analyzed during the current study are available from the corresponding author upon reasonable request. All data needed to evaluate the conclusions in the paper are present in the paper and/or the Supplementary Materials.

## Acknowledgments

The authors thank the participants and personnel of the Knight Alzheimer Disease Research Center, as well as the staff of Washington University’s Translational Human Neurodegenerative Disease Research (THuNDR) Laboratory for providing, processing, staining, and facilitating the digital scanning of the postmortem human brain tissue samples for this study, with support from the National Institutes of Health (NIH)/tNational Institute on Aging (NIA) P01AG026276 (Antecedent Biomarkers of AD: The Adult Children Study, PI J. C. Morris), NIA P01AG003991 (Healthy Aging and Senile Dementia, PI J. C. Morris), NIA P30AG066444 (PI J. C. Morris), and NIA 1R01AG054567-01A1 (PIs T. L. S. Benzinger and Y. Wang). Q.W. is supported by NIA R01AG074909 (PI Q. Wang). National Institutes of Health (R01-AG064614 (CC), U01-AG084514 (CC), P01-NS131131 (CC), R01-AG078964(CC), R01-AG058501 (CC), R01-AG071706 (CC), P30-AG066444 (CC), R01-AG064877 (CC), and the Cure Alzheimer’s Fund.

Additional support was provided by the Charles F. and Joanne Knight Alzheimer’s Research Initiative and by the Fred Simmons and Olga Mohan Fund and the Paula and Rodger Riney Fund.

## Author contributions

Q.W. and Y.W. designed the study. Q.W., M.J., A.L., X.N., and Y.N. performed experiments and data analysis. Q.S. contributed to data processing and proteomics analysis. R.P. and E.F. provided postmortem tissue and neuropathological evaluation. C.C. provided CSF proteomic data. S.F. and T.L.S.B. provided PET imaging data and analysis. Q.W. and Y.W. wrote the manuscript with input from all authors.

## Competing interests

CC is a member of the scientific advisory board of Circular Genomics and owns stocks, and is on the scientific advisory board of ADmit, BMS and Alamar. CC has consulted in the last 6 months for consults for Sanofi, NovoNordisk, and Owkin. CC has received research support from Circular Genomics, GSK, Danaher, BMS, and EISAI. CC is a founder of Andia Health. T.L.S. Benzinger, M.D., Ph.D., has investigator-initiated research funding from the NIH, the Alzheimer’s Association, the Barnes-Jewish Hospital Foundation, and Avid Radiopharmaceuticals; participates as a site investigator in clinical trials sponsored by Avid Radiopharmaceuticals, Eli Lilly and Company, Biogen, Eisai, Jaansen, and F. Hoffmann-La Roche Ltd.; serves as an unpaid consultant to for Eli Lilly, Eisai, Johnson & Johnson, Medscape, PeerView, Neurology Today, Med Learning Group, Applied Radiology, and Siemens, and is on the Speaker’s Bureau for Biogen. All other authors declare they have no competing interests.

## Notes

### Author Declarations

All procedures were approved by the Washington University Institutional Review Board through the Human Research Protections Office and performed in accordance with the Declaration of Helsinki.

