## Supplementary Materials for "Histologically validated diffusion MRI signatures of neuroinflammation and neurodegeneration in Alzheimer disease"

Qing Wang *et al.*

**This PDF file includes:**

Figs. S1 to S8
Tables S1 to S2

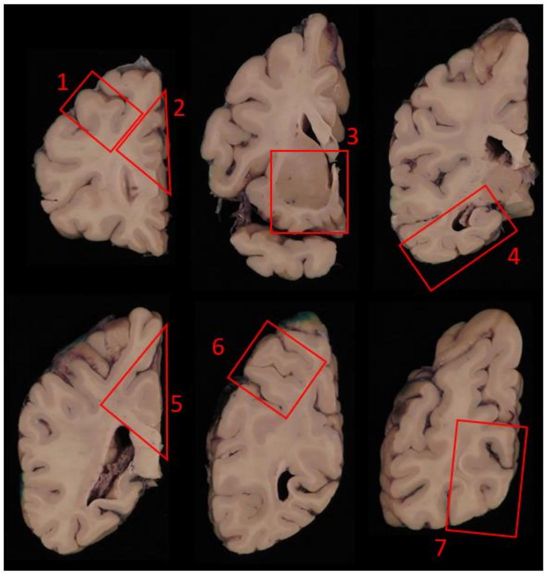


**Fig. S1. Schematic of small tissue block sampling.**

Areas sampled:
1 – Middle frontal gyrus (MFG).
2 – Anterior cingulate gyrus (ACG).
3 – Striatum with nucleus accumbens and olfactory cortex (Striatum).
4 – Hippocampus with parahippocampal, fusiform, inferior temporal gyri (HPC).
5 – Posterior cingulate gyrus and precuneus (PCG/PCu).
6 – Parietal lobe (angular gyrus) (PL).
7 – Occipital lobe (calcarine sulcus and parastriate cortex) (OL).

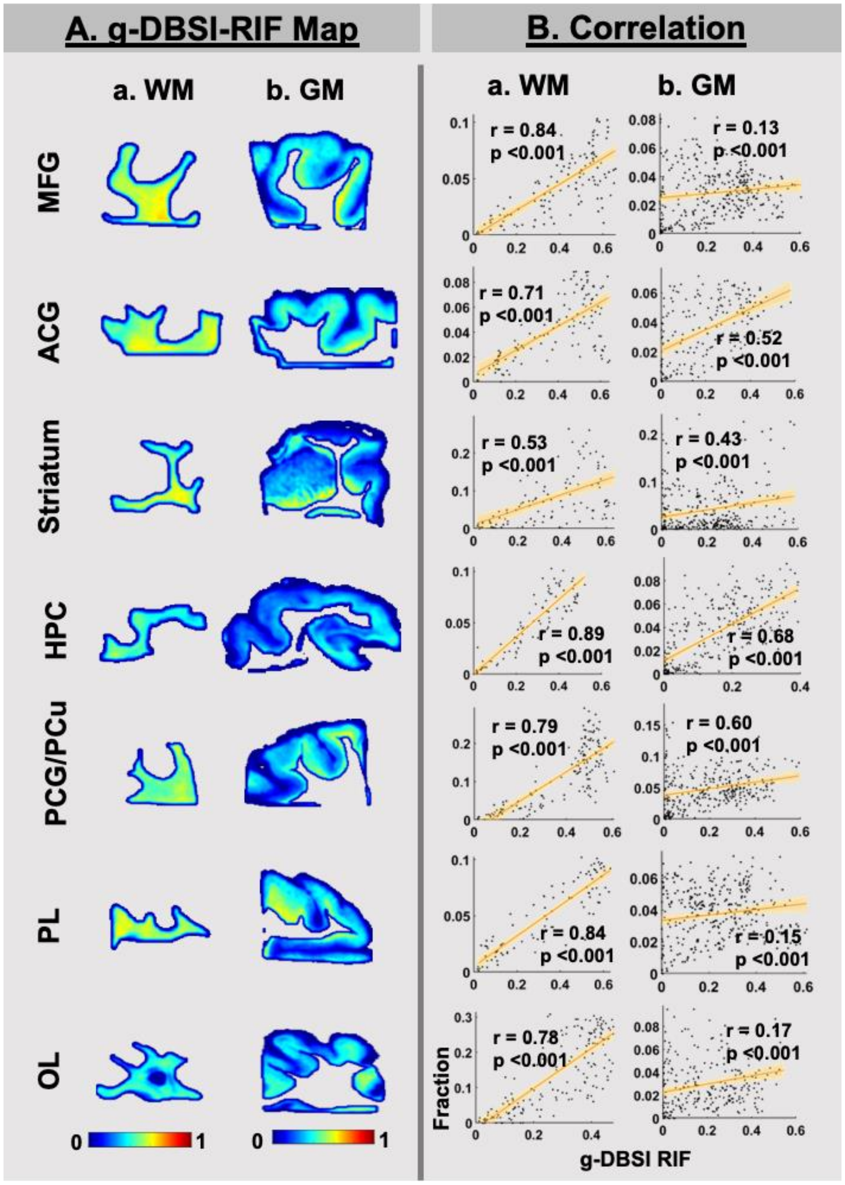


**Fig. S2. Spatial correlations between g-DBSI-derived RIF and H&E cell density across brain regions in white matter and gray matter.** (A) g-DBSI-derived restricted isotropic fraction (RIF) maps segmented into white matter (WM, a) and gray matter (GM, b) compartments for seven brain regions: middle frontal gyrus (MFG), anterior cingulate gyrus (ACG), striatum, hippocampus (HPC), posterior cingulate/precuneus (PCG/PCu), parietal lobe (PL), and occipital lobe (OL). Color bars indicate RIF values ranging from 0 to 1. (B) Region-wise scatter plots of g-DBSI RIF versus H&E-derived cell density in WM (a) and GM (b) for each region. Orange lines indicate least-squares linear regression fits with shaded bands representing 95% confidence intervals. Pearson correlation coefficients (r) and p-values are shown for each region and compartment (all $p < 0.001$). RIF demonstrated consistently strong positive correlations with H&E cell density in WM across all regions (r = 0.53–0.89), with more variable but significant correlations in GM (r = 0.13–0.68).

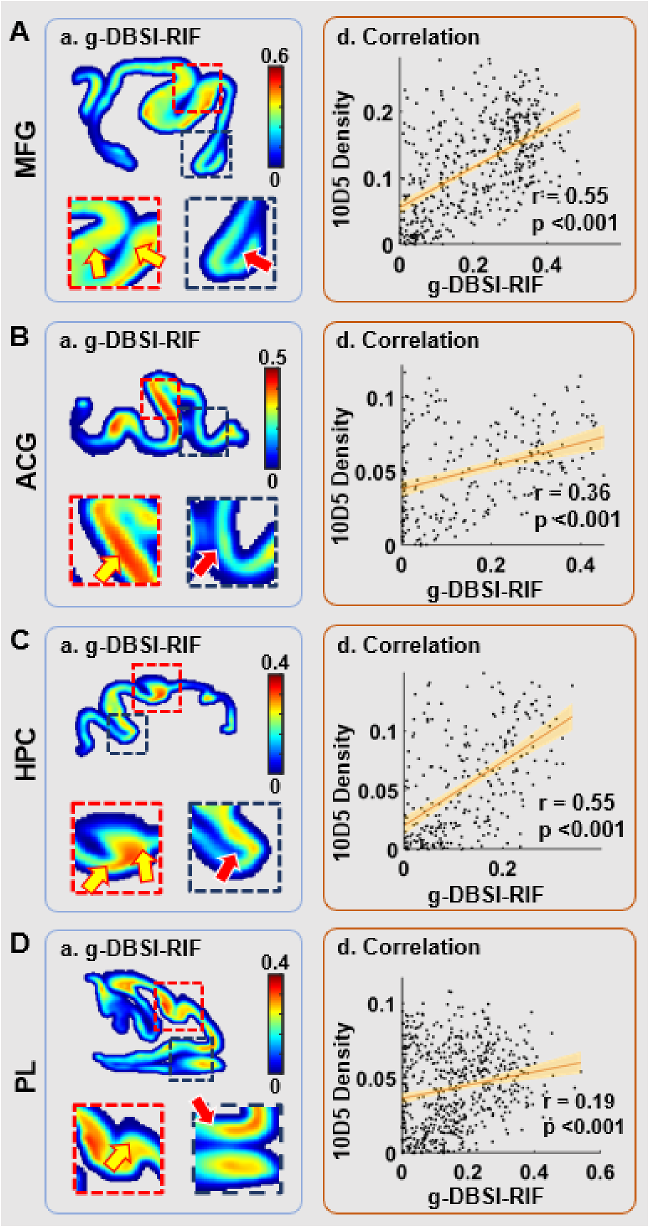


**Fig. S3.** ***Ex situ* g-DBSI RIF spatially correlates with amyloid-beta deposition across cortical regions in postmortem brain tissue blocks.** Representative results from four brain regions: middle frontal gyrus (MFG, A), anterior cingulate gyrus (ACG, B), hippocampus (HPC, C), and parietal lobe (PL, D). For each region: (a) g-DBSI-derived RIF map of the tissue block, with red and blue dashed inset boxes denoting representative subregions shown at higher magnification below each whole-section image; (d) region-wise scatter plot of 10D5 amyloid-beta density versus g-DBSI RIF, with the orange line indicating the least-squares linear regression fit and shaded band representing the 95% confidence interval. Yellow and red arrows indicate areas of spatially concordant elevated signal between 10D5 amyloid-beta density and RIF maps. Color bars indicate RIF values for each region. Pearson correlation coefficients (r) and p-values are reported for each region (d; all $p < 0.001$).

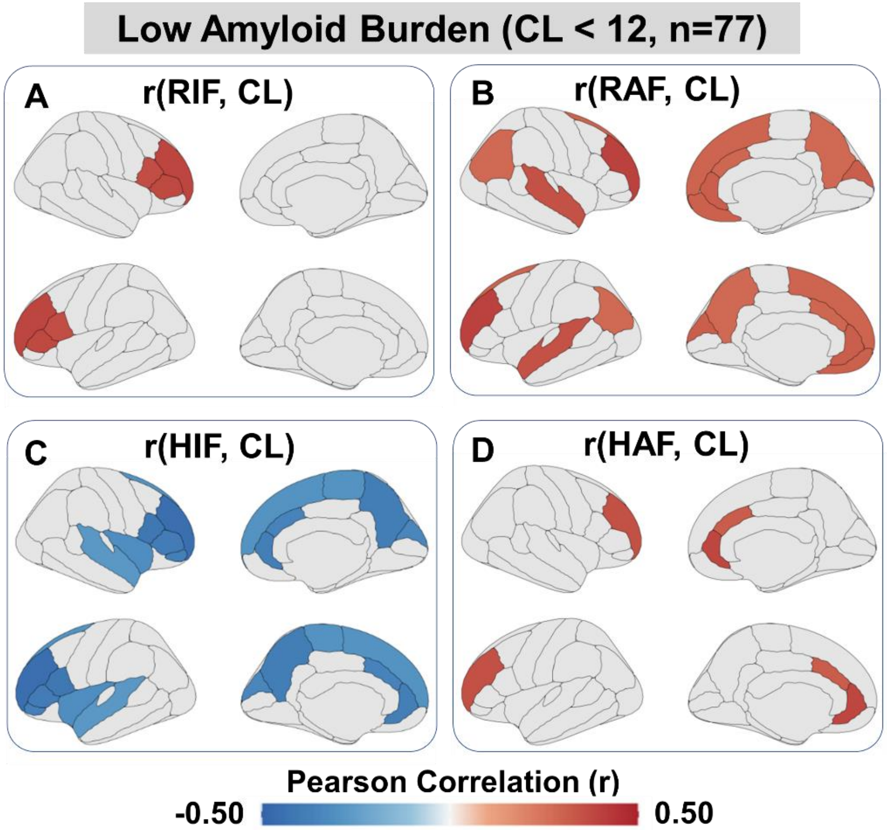


**Fig. S4. Region-wise correlations between cortical g-DBSI indices and PET amyloid burden in the low-amyloid group (CL < 12).** Age- and sex-adjusted partial correlation coefficients (r) between cortical g-DBSI indices of RIF(A), RAF (B), HIF (C), and HAF (D) and amyloid Centiloid (CL) are shown on Desikan-Killiany cortical surface maps in the CL<12 group, with regions surviving FDR correction ($q < 0.05$) shown in color. In the CL<12 group, RAF showed the most spatially extensive positive correlations with CL across frontoparietal, cingulate, and temporal cortices, while HIF exhibited widespread negative correlations over similar regions. RIF and HAF showed more restricted positive correlations in frontal and temporal regions.

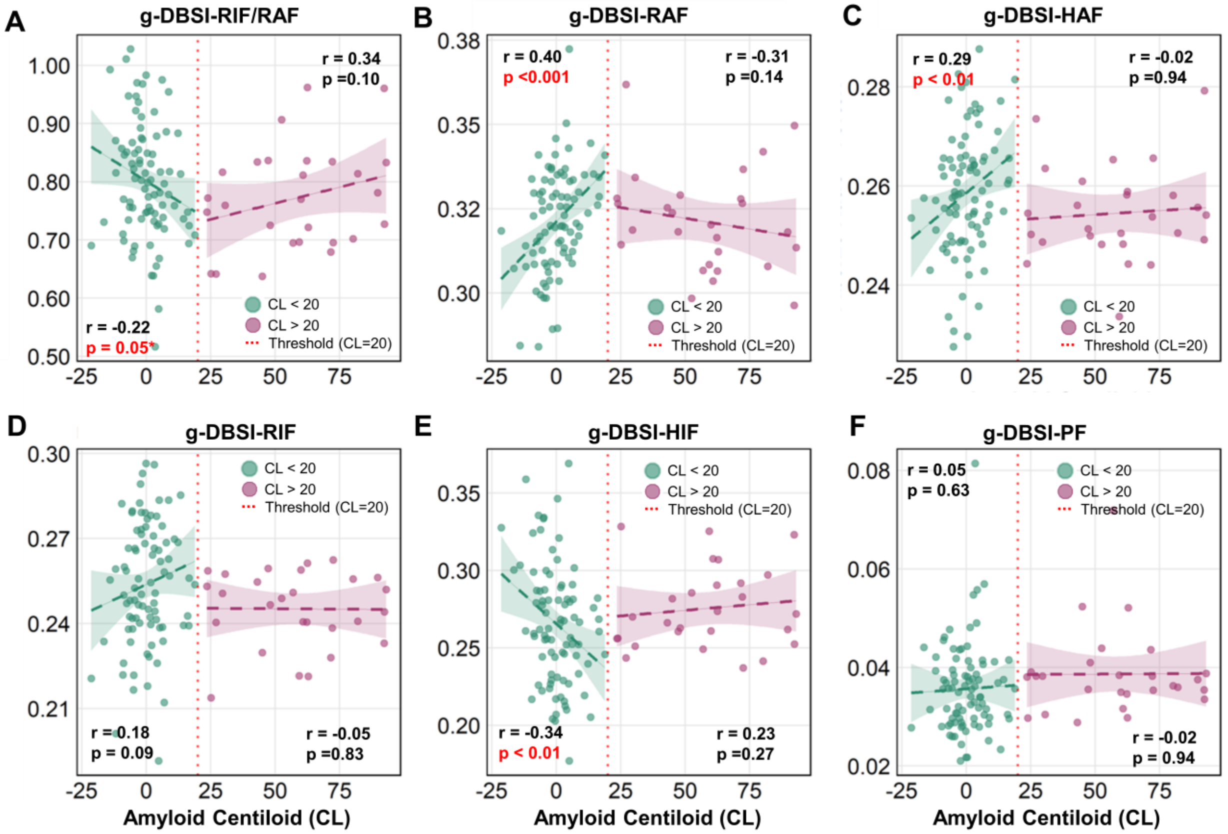


**Fig. S5. Amyloid burden-dependent relationships between cortical microstructural g-DBSI measures and PET amyloid deposition across negative- and positive-amyloid groups.** (A-F) Scatter plots show relationships between the whole-cortical g-DBSI indices and centiloid (CL), quantified by PIB PET, stratified by negative (CL < 20, green) and positive (CL > 20, mulberry) amyloid burden. Solid lines denote least-squares linear regression fits within each group, and shaded backgrounds highlight CL strata separated at the predefined threshold (vertical dashed line at CL = 20). Panels show the ratio of RIF/RAF (A); restricted isotropic fraction (RIF; B), hindered isotropic fraction (HIF; C), restricted anisotropic fraction (RAF; D), hindered anisotropic fraction (HAF; E), and perfusion fraction (PF; F). Pearson correlation coefficients (r) and corresponding p values are indicated for each group; $^*p < 0.05$, $^{**}p < 0.01$. Age and gender were adjusted as covariates.

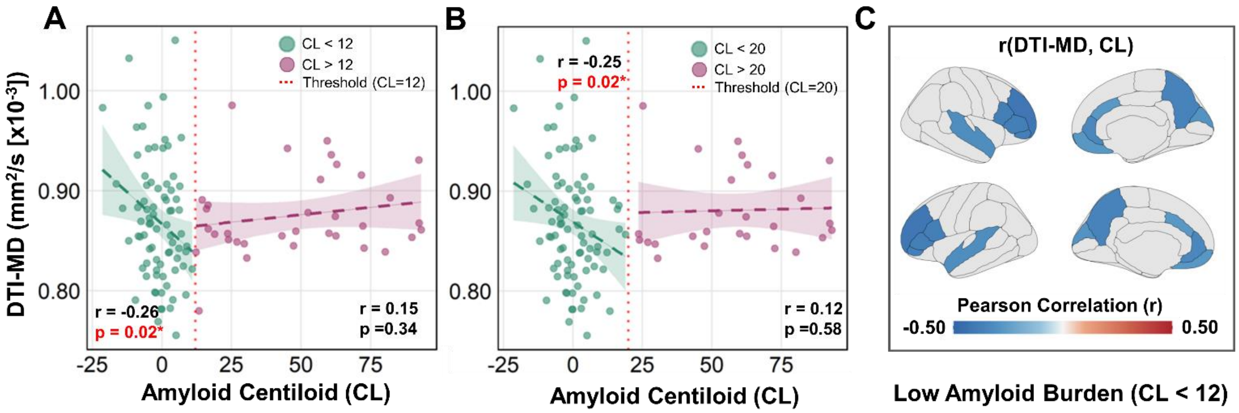


**Fig. S6. Associations between cortical DTI mean diffusivity and PET amyloid deposition.** Scatter plots show age- and sex-adjusted partial correlations between cortical DTI mean diffusivity (MD) and amyloid Centiloid values (CL), stratified by the CL=12 (A) and CL=20 (B) thresholds. Pearson partial correlation coefficients (r) between cortical DTI-MD and CL are shown on Desikan-Killiany cortical surface maps in the CL<12 group, with regions surviving FDR correction ($q < 0.05$) shown in color. Negative correlations between MD and CL were spatially distributed across frontal, parietal, and cingulate cortices. In the scatter plots, green and pink dots represent low- and high-amyloid groups, respectively. Dashed lines indicate linear fits with 95% confidence intervals. Red dotted vertical lines indicate the respective CL thresholds. Pearson correlation coefficients (r) and corresponding p-values are shown for each group; significant correlations (*$p<0.05$) are highlighted in red.

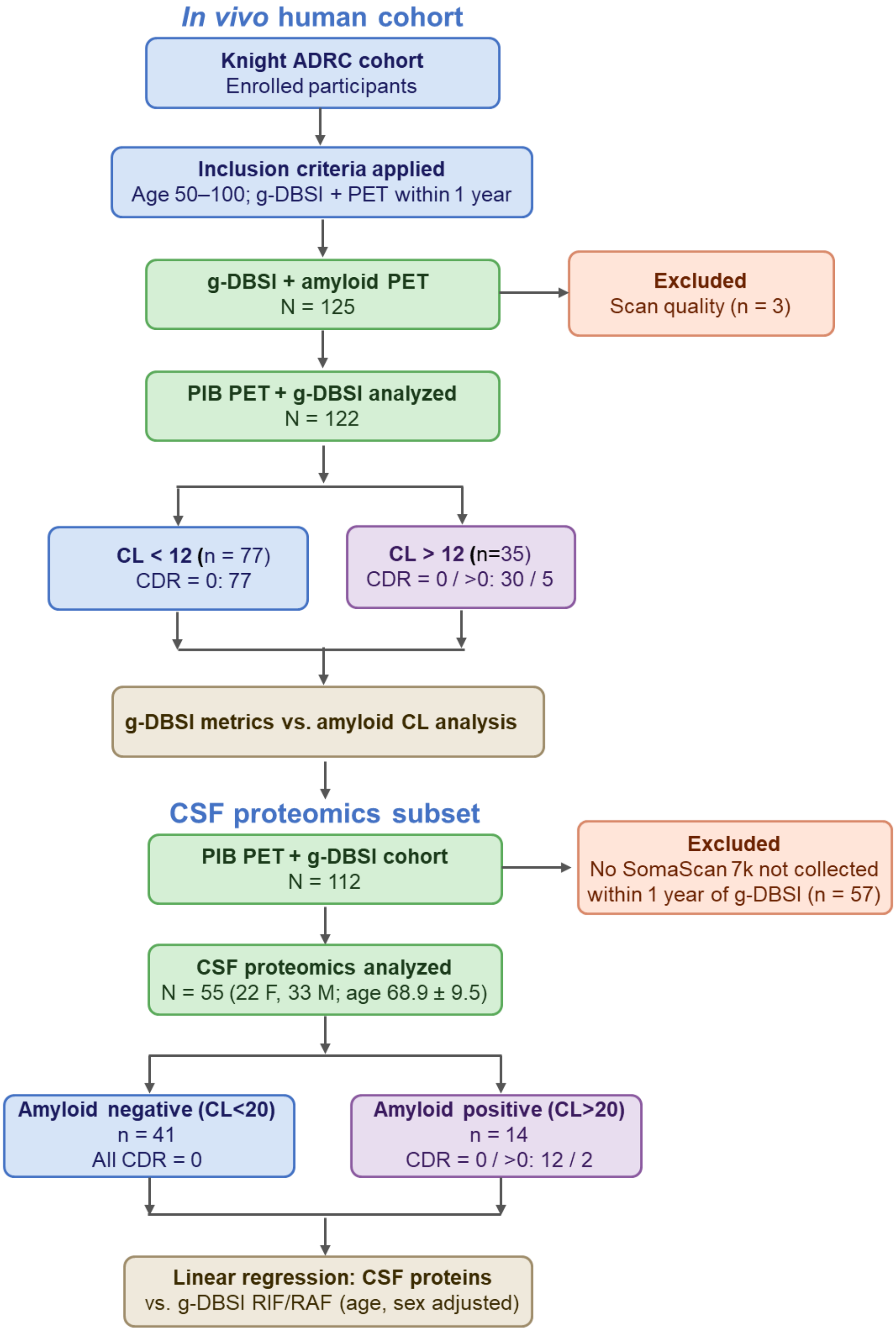


**Fig. S7. Participants flow diagram.**

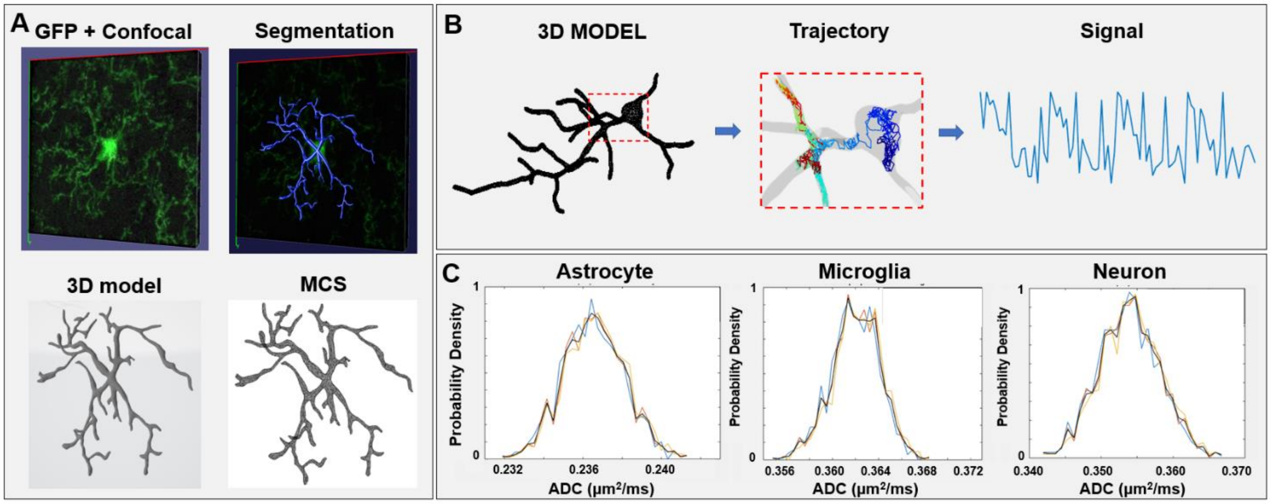


**Fig. S8. Monte Carlo simulation of intracellular diffusion profiles for astrocytes, microglia, and neurons.** (A) Representative GFP confocal 3D image stack of a single cell (top left, green) with semi-automatic segmentation of soma and processes (top right, blue). The segmented morphology was reconstructed into a watertight 3D surface mesh for simulation (bottom left, "3D model") and imported into the Monte Carlo simulator (MCS; bottom right). (B) Restricted diffusion was simulated within the reconstructed 3D cellular volume by tracking random-walk trajectories of water molecules (example trajectories shown in the Zoom in-image, color-coded by displacement magnitude). Synthetic dMRI signals were subsequently generated by applying prescribed diffusion-weighting gradient waveforms across multiple b-values. (C) Probability density distributions of the apparent diffusion coefficient (ADC; $\mu m^2/ms$) derived from Monte Carlo simulations for astrocyte, microglia, and neuron morphologies along x-, y-, and z-axis (blue, orange lines). Each cell type exhibits a distinct ADC distribution, reflecting differences in intracellular geometry, with modest direction-dependent shifts. These simulations provide mechanistic validation linking cell-type-specific morphological architecture to the diffusion signal components modeled in g-DBSI.

**Table S1. Summarized characteristics of participants in the ex vivo study**

| ID | Sex | Age | Diagnosis | Thal Stage | Braak Stage | CDR | PMI (h) | APOE |
|---|---|---|---|---|---|---|---|---|
| #1 | Male | 81-85 | AD Dementia | 5 | IV | 1 | 18.4 | 34 |
| #2 | Male | 86-90 | AD Dementia | 5 | V | 3 | 4.3 | 34 |
| #3 | Female | 91-95 | AD Dementia | 3 | V | 3 | 7 | 33 |
| #4 | Female | 71-75 | AD Dementia | 5 | V | 1 | 8.1 | 34 |
| #5 | Female | 81-85 | Normal Cognition | 5 | V | 0 | 3.8 | 33 |
| #6 | Male | 61-65 | Normal Cognition | 1 | II | 0 | 17.5 | 34 |

**Table S2. Demographic and Clinical Characteristics of Participants with CSF Proteomics**

| Variable | A- | A+ | P |
|---|---|---|---|
| CDR=0/CDR>0 | 41/0 | 12/2 | 0.061 |
| age | 68.55 ± 8.85 | 74.31 ± 8.78 | **0.046** |
| education | 16.29 ± 2.21 | 15.86 ± 1.99 | 0.499 |
| Female (% no.) | 25 (61.0%) | 5 (35.7%) | 0.128 |
| APOE ε4 (% no.) | 12 (29.3%) | 8 (57.1%) | 0.106 |
| CDR-SB | 0.01 ± 0.08 | 0.46 ± 1.18 | 0.177 |
| MMSE | 29.02 ± 1.39 | 27.86 ± 2.41 | 0.106 |

Data are presented as mean ± SD for continuous variables and n (%) for categorical variables. P-values are derived from raw pairwise comparisons (Kruskal–Wallis and Chi-square tests). Abbreviations: APOE ε4, apolipoprotein E gene; CDR, clinical dementia rating; CDR-SB, CDR-sum of boxes; MMSE, mini-mental state examination.
